# Using viral load and epidemic dynamics to optimize pooled testing in resource constrained settings

**DOI:** 10.1101/2020.05.01.20086801

**Authors:** Brian Cleary, James A. Hay, Brendan Blumenstiel, Maegan Harden, Michelle Cipicchio, Jon Bezney, Brooke Simonton, David Hong, Madikay Senghore, Abdul K. Sesay, Stacey Gabriel, Aviv Regev, Michael J. Mina

**Affiliations:** Broad Institute of MIT and Harvard, Cambridge, MA 02142 USA; Centre for Communicable Disease Dynamics, Department of Epidemiology, Harvard School of Public Health, Boston, MA, USA; Department of Immunology and Infectious Diseases, Harvard School of Public Health; Wharton Statistics, University of Pennsylvania, Philadelphia, PA, USA; Medical Research Council Unit The Gambia at London School of Hygiene and Tropical Medicine, PO Box 273, Banjul, The Gambia; Klarman Cell Observatory, Broad Institute of MIT and Harvard; Department of Biology, Massachusetts Institute of Technology, Cambridge, MA 02142 USA; Howard Hughes Medical Institute, Chevy Chase, MD, USA; Genentech, 1 DNA Way, South San Francisco, CA, USA; Department of Pathology, Brigham and Women’s Hospital, Harvard Medical School

## Abstract

Extensive virological testing is central to SARS-CoV-2 containment, but many settings face severe limitations on testing. Group testing offers a way to increase throughput by testing pools of combined samples; however, most proposed designs have not yet addressed key concerns over sensitivity loss and implementation feasibility. Here, we combine a mathematical model of epidemic spread and empirically derived viral kinetics for SARS-CoV-2 infections to identify pooling designs that are robust to changes in prevalence, and to ratify losses in sensitivity against the time course of individual infections. Using this framework, we show that prevalence can be accurately estimated across four orders of magnitude using only a few dozen pooled tests without the need for individual identification. We then exhaustively evaluate the ability of different pooling designs to maximize the number of detected infections under various resource constraints, finding that simple pooling designs can identify up to 20 times as many positives compared to individual testing with a given budget. We illustrate how pooling affects sensitivity and overall detection capacity during an epidemic and on each day post infection, finding that sensitivity loss is mainly attributed to individuals sampled at the end of infection when detection for public health containment has minimal benefit. Crucially, we confirm that our theoretical results can be accurately translated into practice using pooled human nasopharyngeal specimens. Our results show that accounting for variation in sampled viral loads provides a nuanced picture of how pooling affects sensitivity to detect epidemiologically relevant infections. Using simple, practical group testing designs can vastly increase surveillance capabilities in resource-limited settings.

## Introduction

The ongoing pandemic of SARS-CoV-2, a novel coronavirus, has caused over 83 million reported cases of coronavirus disease 2019 (COVID-19) and 1.8 million reported deaths between December 2019 and January 2021. (1) Although wide-spread virological testing is essential to inform disease status and where outbreak mitigation measures should be targeted or lifted, sufficient testing of populations with meaningful coverage has proven difficult. (2–7) Disruptions in the global supply chains for testing reagents and supplies, as well as on-the-ground limitations in testing throughput and financial support, restrict the usefulness of testing–both for identifying infected individuals and to measure community prevalence and epidemic trajectory. While these issues have been at the fore in even the highest-income countries, the situation is even more dire in low-income regions of the world. Cost barriers alone mean it is often simply not practical to prioritize community testing in any useful way, with the limited testing that exists necessarily reserved for the healthcare setting. These limitations urge new, more efficient, approaches to testing to be developed and adopted both for individual diagnostics and to enable public health epidemic control and containment efforts.

Group or pooled testing offers a way to increase efficiency by combining samples into a group or pool and testing a small number of pools rather than all samples individually. (8–10) For classifying individual samples, including for diagnostic testing, the principle is simple: if a pool tests negative, then all of the constituent samples are assumed negative. If a pool tests positive, then the constituent samples are putatively positive and must be tested again individually or in mini-pools (**Fig. 1A**). Further efficiency gains are possible through combinatorial pooling, where, instead of testing every sample in every positive pool, each sample can instead be represented across multiple pools and potential positives are identified based on the pattern of pooled results (**Fig. 1B**). (9, 10)

**Fig. 1.**
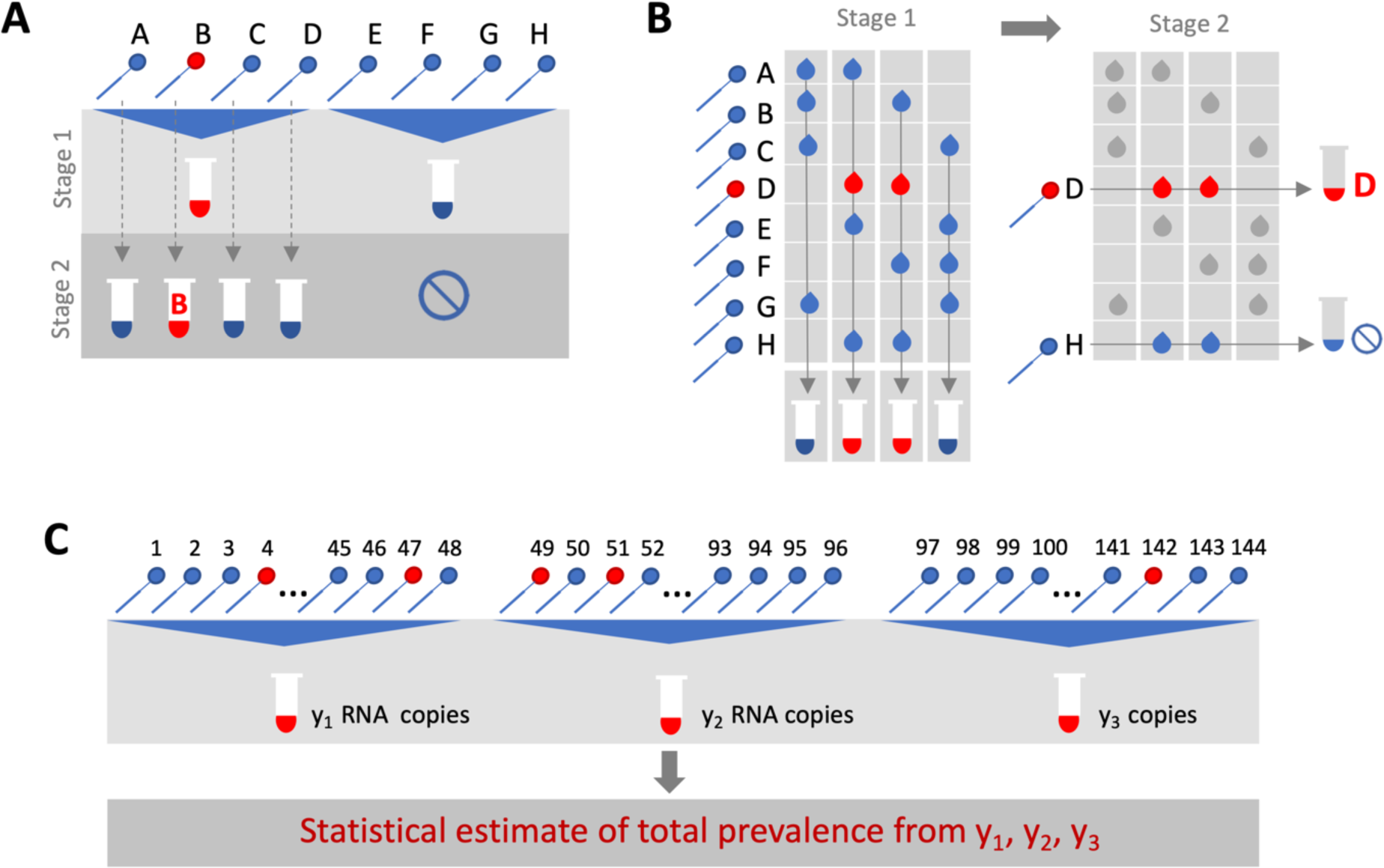
Group testing designs for sample identification or prevalence estimation. In group testing, multiple samples are pooled and tests are run on one or more pools. The results of these tests can be used for identification of positive samples (A, B) or to estimate prevalence (C). (A) In the simplest design for sample identification, samples are partitioned into non-overlapping pools. In stage 1 of testing, a negative result (Pool 2) indicates each sample in that pool was negative, while a positive result (Pool 1) indicates at least one sample in the pool was positive. These putatively positive samples are subsequently individually tested in stage 2 to identify positive results. (B) In a combinatorial design, samples are included in multiple pools as shown in stage 1. All samples that were included in negative pools are identified as negative, and the remaining putatively positive samples that were not included in any negative test are tested individually in stage 2. (C) In prevalence estimation, samples are partitioned into pools. The pool measurement will depend on the number and viral load of positive samples, and the dilution factor. The (quantitative) results from each pool can be used to estimate the fraction of samples that would have tested positive, had they been tested individually.

Simple pooling designs can also be used to assess prevalence without individual specimen identification (**Fig. 1C**). It has already been shown that the frequency of positive pools can allow estimation of the overall prevalence. (11) However, we ask here if prevalence estimates can be honed by considering quantitative viral loads measured in each positive pool, rather than simply using binary results (positive / negative), where the viral (RNA) load measurement from a pool is proportional to the sum of the (diluted) viral loads from each positive sample in the pool. An extreme, albeit less precise, example of this is the quantitation of viral loads in wastewater as a metric for whole community prevalence. (12)

Whilst the literature on theoretically optimized pooling designs for COVID-19 testing has grown rapidly, formal incorporation of biological variation (*i.e.,* viral loads) and incorporation of general position along the epidemic curve, has received little attention. (13–16) Crucially, test sensitivity is not a fixed value, but depends on viral load, which can vary by many orders of magnitudes across individuals and over the course of an infection, with implications for appropriate intervention and the interpretation of a viral load measurement from a sample pool. (17–19) Further, the distribution of viral loads in surveillance testing is also sensitive to the course in the epidemic (growth versus decay) which will thus also affect the measured test sensitivity. (20)

Here, we comprehensively evaluate designs for pooled testing of SARS-CoV-2 whilst accounting for epidemic dynamics and variation in viral loads arising from viral kinetics and extraneous features such as sampling variation. We demonstrate efficient, logistically feasible pooling designs for individual identification (*i.e.,* diagnostics) and prevalence estimation (*i.e.,* population surveillance). To do this, we use realistic simulated viral load data at the individual level over time, representing the entire time course of an epidemic to generate synthetic data that reflects the true distribution of viral loads in the population at any given time of the epidemic. We then used these data to derive optimal pooling strategies for different use cases and resource constraints *in-silico*. In particular, we show how evaluating viral loads provides substantial efficiency gains in prevalence estimates, enabling robust public health surveillance where it was previously out of reach. Finally, we demonstrate the approach using discarded de-identified human nasopharyngeal swabs initially collected for diagnostic and surveillance purposes.

## Results

### Modelling a synthetic population to assess pooling designs

To identify optimal pooling strategies for distinct scenarios, we required realistic estimates of viral loads across epidemic trajectories. We developed a population-level mathematical model of SARS-CoV-2 transmission that incorporates empirically measured within-host virus kinetics, and used these simulations to generate population-level viral load distributions representing real data sampled from population surveillance, either using nasopharyngeal swab or sputum samples (**Fig. 2**, **Materials and Methods**, **Supplementary Material 1, sections 1-4**). These simulations generated a synthetic, realistic epidemic with a peak daily per incidence of 19.5 per 1,000 people, and peak daily prevalence of RNA positivity (viral load greater than 100 virus RNA copies per ml) of 265 per 1,000 (**Fig. 2D**). We used these simulation data to evaluate optimal group testing strategies at different points along the epidemic curve for diagnostic as well as public health surveillance, where the true viral loads in the population is known fully.

**Fig. 2.**
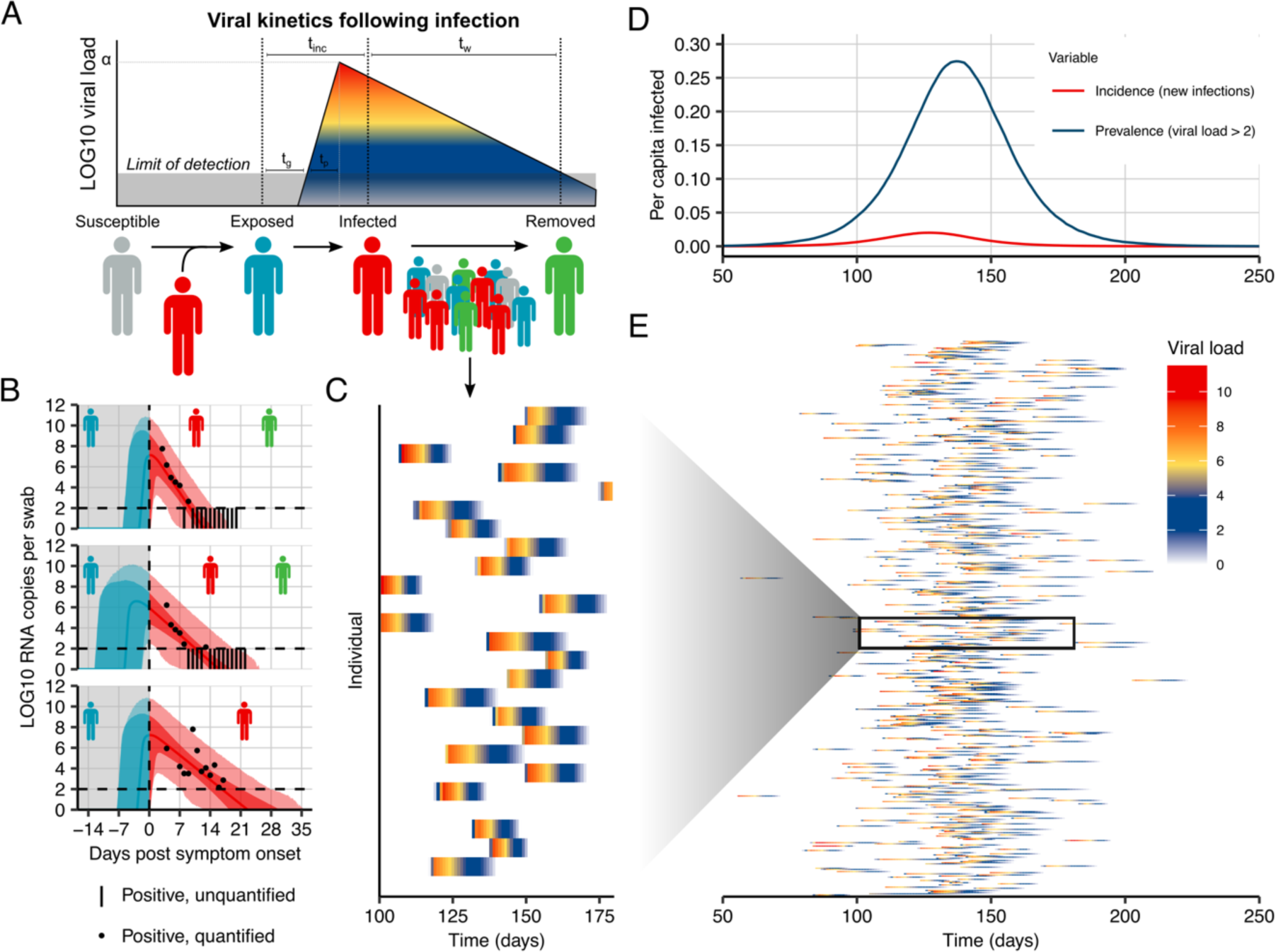
Viral kinetics model fits, simulated infection dynamics and population-wide viral load kinetics. (A) Schematic of the viral kinetics and infection model. Individuals begin susceptible with no viral load (susceptible), acquire the virus from another infectious individual to become exposed but not yet infectious (exposed), experience an increase in viral load to become infectious and possibly develop symptoms (infected), and finally either recover following viral waning or die (removed). This process is simulated for many individuals. (B) Model fits to time-varying viral loads in swab samples. The black dots show observed *log10* RNA copies per swab; black bars show positive but unquantified swab samples; solid lines show posterior median estimates; dark shaded regions show 95% credible intervals (CI) on model-predicted latent viral loads; light shaded regions show 95% CI on simulated viral loads with added observation noise. The blue region shows viral loads before symptom onset and red region shows time after symptom onset. The horizontal dashed line shows the limit of detection. (C, E) 25 and 500 simulated viral loads over time, respectively. The heatmap shows the viral load in each individual over time. The distribution of viral loads reflects the increase and subsequent decline of prevalence. We simulated from inferred distributions for the viral load parameters, thereby propagating substantial individual-level variability through the simulations. Marginal distributions of observed viral loads during the different epidemic phases are shown in fig. S4. (D) Simulated infection incidence and prevalence of virologically positive individuals from the SEIR model. Incidence was defined as the number of new infections per day divided by the population size. Prevalence was defined as the number of individuals with viral load > 100 (*log10* viral load > 2) in the population divided by the population size on a given day.

### Improved testing efficiency for estimating prevalence

We developed a statistical method to estimate prevalence of SARS-CoV-2 based on cycle threshold (Ct) values measured from pooled samples (**Materials and Methods**), potentially using far fewer tests than would be required to assess prevalence based on number of positive samples identified. We used our synthetic viral load data to assess inferential accuracy under a range of sample availabilities and pooling designs. Since RNA extraction and PCR efficiency can vary from lab to lab depending on the methods used, and within lab from batch to batch, we introduced substantial variability in our simulations in the conversion from viral load to Ct value to capture the multiple levels of uncertainty (**Supplementary Material Section 5**). In practice, this uncertainty can be refined by focusing on a single assay within a single lab.

Across the spectrum of simulated pools and tests we found that simple pooling allows accurate estimates of prevalence across at least four orders of magnitude, ranging from 0.02% to 20%, with up to 400- times efficiency gains (*i.e.,* 400 times fewer tests) for prevalence estimation than would be needed without pooling (**Fig. 3**). For example, in a population prevalence study that collects ∼2,000 samples, we accurately estimated infection prevalences as low as 0.05% by using only 24 qPCR tests overall (*i.e.*, 24 pools of 96 samples each; **Fig. 3A**; **fig. S1**). Importantly, because the distribution of Ct values may differ depending on the sample type (sputum *vs*. swab), the instrument, and the phase of the epidemic (growth vs. decline, **fig. S2**), the method should be calibrated in practice to viral load data (*i.e.,* Ct values) specific to the laboratory and instrument (which can differ between labs) and the population under investigation.

**Fig. 3.**
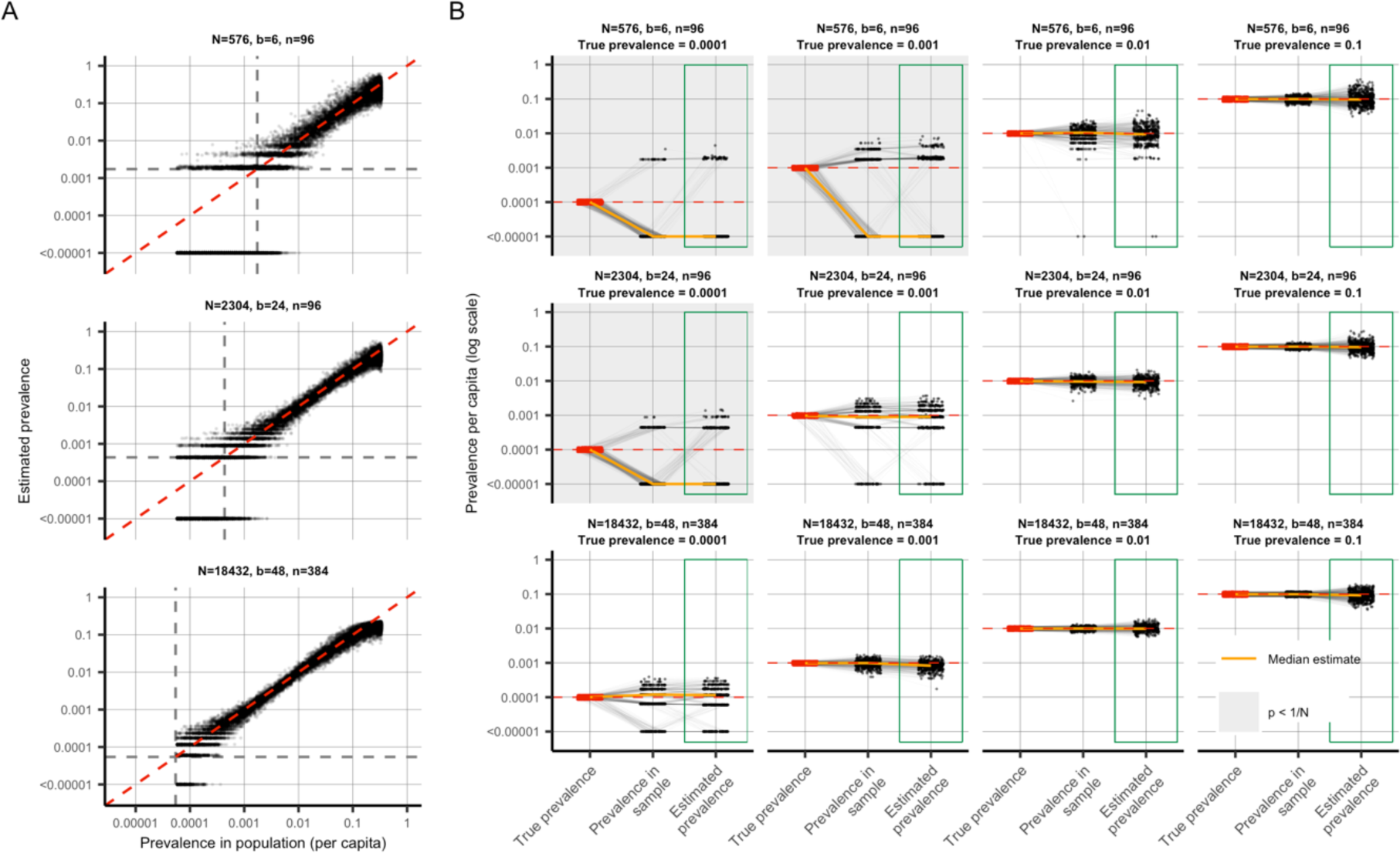
Estimating prevalence from a small number of pooled tests. In prevalence estimation, a total of *N* individuals are sampled and partitioned into *b* pools (with *n = N/b* samples per pool). The true prevalence in the entire population varies over time with epidemic spread. Population prevalences shown here are during the epidemic growth phase. (A) Estimated prevalence (y axis) and true population prevalence (x axis) using 100 independent trials sampling N individuals at each day of the epidemic. Each facet shows a different pooling design (additional pooling designs shown in fig. S1). Dashed grey lines: one divided by the sample size, N. (B) For a given true prevalence (label on top, red points and horizontal dashed red line), estimation error is introduced both through binomial sampling of positive samples (Prevalence in sample) and inference on the sampled viral loads (Estimated prevalence, green boxes). Each set of three connected dots shows one simulation, with points slightly jittered on the x-axis for visibility. Horizontal lines indicate accurate inference. The orange line shows the median across 100 simulations. Each panel shows the results from a single pooling design at the specified true prevalence. Sampling variation is a bigger contributor to error at low prevalence and low sample sizes. When prevalence is less than one divided by *N* (grey shaded panels), inference is less accurate due to the high probability of sampling only negative individuals or inclusion of false positives.

Estimation error arises in two stages: sample collection effects, and as part of the inference method (**Fig. 3B**). Error from sample collection became less important with increasing numbers of positive samples, which occurred with increasing population prevalence or by increasing the total number of tested samples (**Fig. 3B**; **fig. S2**). At very low prevalence, small sample sizes (*N*) risk missing positives altogether or becoming biased by false positives. We found that accuracy in prevalence estimation is greatest when population prevalence is greater than 1/N and that when this condition was met, partitioning samples into more pools always improves accuracy (**fig. S2**). In summary, very accurate estimates of prevalence can be attained using only a small fraction of the tests that would be needed in the absence of pooling.

### Pooled testing for individual identification

We next analyzed effectiveness of group testing for identifying individual sample results at different points along the epidemic curve with the aim of identifying simple, efficient pooling strategies that are robust across a range of prevalences (**Fig. 1A,B**). Using the simulated viral load data (**Materials and Methods**), we evaluated a large array of pooling designs *in silico* (**table S1**). Based on our models of viral kinetics and given a PCR limit of detection of 100 viral copies per ml, we first estimated a baseline sensitivity of conventional (non-pooled) PCR testing of 85% during the epidemic growth phase (*i.e.*, 15% of the time we sample an infected individual with a viral load greater than 1 but below the LOD of 100 viral copies per mL, **Fig. 4A**), which largely agrees with reported estimates. (21, 22) This reflects sampling during the latent period of the virus (after infection but prior to significant viral growth) or in the relatively long duration of low viral titers during viral clearance.

**Fig. 4.**
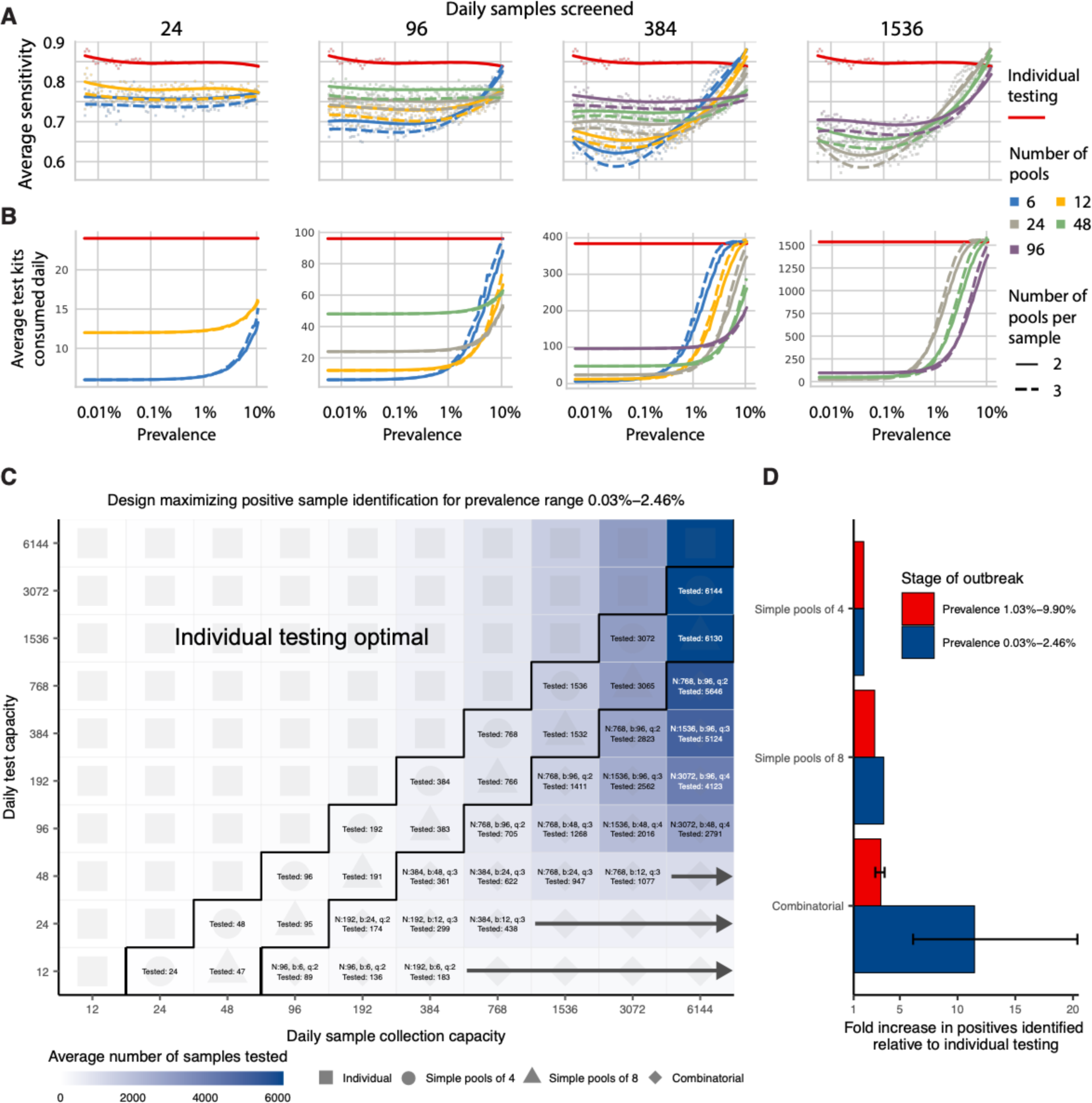
Group testing for sample identification. We evaluated a variety of group testing designs for sample identification (**table S1**) on the basis of sensitivity **(A)**, efficiency **(B)**, total number of positive samples identified **(C)** and the fold increase in positive samples identified relative to individual testing **(D)**. (**A** and **B**) The average sensitivity (**A**, y-axis, individual points and spline) and average number of tests needed to identify individual positive samples (**B**, y-axis) using different pooling designs (individual lines) were measured over days 20-110 in our simulated population, with results plotted against prevalence (x-axis, log-scale). Results show the average of 200,000 trials, with individuals selected at random on each day in each trial. Pooling designs are separated by the number of samples tested on a daily basis (individual panels); the number of pools (color); and the number of pools into which each sample is split (dashed versus solid line). Solid red line indicates results for individual testing. Note that the average number of test kits consumed increases with prevalence due to a greater number of positive pools requiring confirmatory testing. **(C)** Every design was evaluated under constraints on the maximum number of samples collected (columns) and average number of reactions that can be run on a daily basis (rows) over days 40-90. Text in each box indicates the optimal design for a given set of constraints (number of samples per batch (N), number of pools (b), number of pools into which each sample is split (q), average number of total samples screened per day). Color indicates the average number of samples screened on a daily basis using the optimal design. Arrows indicate that the same pooling design is optimal at higher sample collection capacities due to testing constraints. **(D)** Fold increase in the number of positive samples identified relative to individual testing with the same resource constraints. Error bar shows range amongst optimal designs.

Sensitivity of pooled tests, relative to individual testing, is affected by the dilution factor of pooling and by the population prevalence – with lower prevalence resulting in generally lower sensitivity as positives are diluted into many negatives (**Fig. 4A**). The decrease in sensitivity is roughly linear with the log of the dilution factor employed, which largely depends on the number and size of the pools and, for combinatorial pooling, the number of pools that each sample is placed into (**fig. S3A-C**).

There is a less intuitive relationship between sensitivity and prevalence as it changes over the course of the epidemic. Early in an epidemic there is an initial dip in sensitivity for both individual and pooled testing (**Fig. 4A**). Early on, during exponential growth of an outbreak, in a random sample of infected individuals, a relatively greater fraction of positives will be sampled early in their infection and thus closer to their peak viral load. Later on, there is an increasing mixture of newly infected with individuals at the tail end of their infections, and thus with lower viral loads at the time of sampling. We found that as a result, sensitivity of pooled testing increases at peak prevalence because samples with lower viral loads, which would otherwise be missed due to dilution, are more likely to be ‘rescued’ by coexisting in the same pool with high viral load samples and thus ultimately get individually retested (at their undiluted or less diluted concentration) during the validation stage. During epidemic decline, fewer new infections arise over time and therefore a randomly selected infected individual is more likely to be sampled during the recovery phase of their infection, when viral loads are lower (**fig. S4D**). Overall sensitivity is therefore lower during epidemic decline, as more infected individuals have viral loads below the limit of detection. During epidemic growth (*i.e.*, up to day 108 in the simulation), we found that overall sensitivity of RT-PCR for individual testing was 85%, whereas during epidemic decline (from day 168 onward) it was 60% (**fig. S5A**). Mean sensitivity of RT-PCR for individual testing was ∼75% across the whole epidemic. We note that in practice, sensitivity is likely higher than estimated here, because individuals are not sampled entirely at random but instead tend to be enriched with symptomatic people sampled nearer to peak viral loads. Together, these results describe how sensitivity is affected by the combination of epidemic dynamics, viral kinetics, and pooling design when individuals are sampled randomly from the population.

Importantly, we find that on average most false negatives arise from individuals sampled seven days or more after their peak viral loads, or around seven days after what is normally considered symptom onset (∼75% in swab samples during epidemic growth; ∼96% in swab samples during epidemic decline). This reflects that the majority of time spent in the PCR positive state is usually post-infectious, and the asymmetry highlights differences in prevalence of new versus older infections during epidemic growth versus decay. Importantly, only ∼3% of false negative swab samples arose from individuals tested during the first week following peak viral load during epidemic growth, and only ∼1% during epidemic decline – (peak titers usually coincide with symptom onset) – and thus most false negatives are from individuals with the least risk of onward transmission (**fig. S3D,E**).

The lower sensitivity of dilutional pooled testing is counterbalanced by gains in efficiency. When prevalence is low, efficiency is roughly the number of samples divided by the number of pools, since there are rarely putative positives to test individually. However, the number of validation tests required will increase as prevalence increases, and designs that are initially more efficient will lose efficiency (**Fig. 4B**). In general, we find that at very low population prevalence the use of fewer pools each with larger numbers of specimens offers relative efficiency gains compared to larger numbers of smaller pools, as the majority of pools will test negative. However, as prevalence increases, testing a greater number of smaller pools pays off as more validations will be performed on fewer samples overall (**Fig. 4B**). For combinatorial designs with a given number of total samples and pools, splitting each sample across fewer pools results in modest efficiency gains (dashed versus solid lines in **Fig. 4B**).

To address realistic resource constraints, we integrated our analyses of sensitivity and efficiency with limits on daily sample collection and testing capacity to maximize the number of positive individuals identified (**Materials and Methods**). We analyzed the total number of samples screened and the fold increase in the number of positive samples identified relative to individual testing for a wide array of pooling designs evaluated over a period of 50 days during epidemic spread (days 40-90 where point prevalence reaches ∼2.5%; **Fig. 4C,D**). Because prevalence changes over time, the number of validation tests may vary each day despite constant pooling strategies. Thus, tests saved on days requiring fewer validation tests can be stored for days where more validation tests are required.

Across all resource constraints considered, we found that effectiveness ranged from one (when testing every sample individually is optimal) to 20 (*i.e.*, identifying 20x more positive samples on a daily basis compared with individual testing within the same budget; **Fig. 4D**). As expected, when capacity to collect samples exceeds capacity to test, group testing becomes increasingly effective. Simple pooling designs are most effective when samples are in moderate excess of testing capacity (2-8x), whereas we find that increasingly complex combinatorial designs become the most effective when the number of samples to be tested greatly exceeds testing capacity. Additionally, when prevalence is higher (*i.e.,* sample prevalence from 1.03% to 9.90%), the optimal pooling designs shift towards combinatorial pooling, and the overall effectiveness decreases – but still remains up to 4x more effective than individual testing (**fig. S6**). Our results were qualitatively unchanged when evaluating the effectiveness of pooling sputum samples, and the optimal pooling designs under each set of sample constraints were either the same or very similar (**fig. S7**). Furthermore, we evaluated the same strategies during a 50-day window of epidemic decline (days 190-250) and found that similar pooling strategies were optimally effective, despite lower overall sensitivity as described above (**fig. S5**).

### Pooled testing in a sustained, multi-wave epidemic

Modeling the time evolution of viral load distributions and prevalence in sustained or multiple epidemic phases is important to understanding realistic performance of pooled testing for SARS-CoV-2 and other respiratory viral diseases. Building on our results above, we next simulated an epidemic with an initial wave, followed by a decline phase and subsequently another growth phase (**fig. S8A; Materials and Methods and Supplementary Material 1, section 1**).

Using this simulated epidemic, we first assessed how pooled testing for prevalence estimation would be affected by multiple waves. Since the results above (**fig. S2**) demonstrate that it is best to match training data for calibrating viral load distributions with the same phase in which the distributions will be used to estimate prevalence (*i.e.*, match training data from growth phase with application during growth phase, and decline with decline), we asked if viral loads observed during a first wave of epidemic growth are appropriate training data for prevalence estimation during a second growth phase. We found that models using either training phase have low, nearly identical, levels of errors when predicting prevalence in the second growth phase (**fig. S8B**).

We next evaluated pooled testing for sample identification under the two-wave epidemic model. Our results above demonstrated a difference in sensitivity between growth and decline in a single-wave epidemic, driven by a shift in the viral load distribution sampled on any given day, which is in turn driven by viral kinetics and a shifting bias in the time relative to infection at which individuals are sampled (20). In an epidemic model with two waves there is a dip in sensitivity over the transition from the first growth to decline phase, followed by a rise in sensitivity during the second growth phase (**fig. S9A**). The reduction in sensitivity for pooled testing compared to individual testing is relatively consistent across epidemic time for any given pooling design, demonstrating robustness to more complicated epidemic dynamics. Moreover, the changes in sensitivity are quantitative reflections of growth rate, so that the lower growth rate of the second growth phase results in a lower sensitivity than the first phase, due to more samples being from individuals with lower viral loads (**fig. S9A**).

The model also allows us to assess any impacts on sample identification resulting from different epidemic dynamics in each period, *e.g.*, during initial epidemic growth with a high basic reproductive number (2.5) and a large susceptible population, compared with a second growth phase with smaller basic reproductive number (1.5) and a smaller susceptible population (due to past infections). We find that pooled testing for sample identification remains advantageous throughout the modeled time series, with very similar optimal pooling strategies under the same array of resource constraints considered above (**fig. S9B,C**).

Together, these results demonstrate generalizability to sustained or multiple wave epidemics, and robustness to changes in viral load distributions from different epidemic growth rates.

### Pilot and validation experiments

We validated our pooling strategies using anonymized clinical nasopharyngeal swab specimens. To evaluate simple pooling across a range of inputs, we diluted 5 nasopharyngeal clinical swab samples with viral loads of 89000, 12300, 1280, 140 and 11 viral RNA copies per ml, respectively, into 23 negative nasopharyngeal swab samples (pools of 24) (**Materials and Methods, Supplementary Material 1, sections 9&10**). The results matched the simulated sampling results: the first three pools were all positive, the fourth was inconclusive (negative on N1, positive on N2), and the remaining pool was negative (Fig. 5A, **table S2**). These results are as expected because the EUA approved assay used has a limit of detection of ∼100 virus copies per ml, such that the last two specimens fall below the limit of detection given a dilution factor of 24 (i.e. 0.46 and 5.8 virus copies per ml once pooled).

**Fig. 5.**
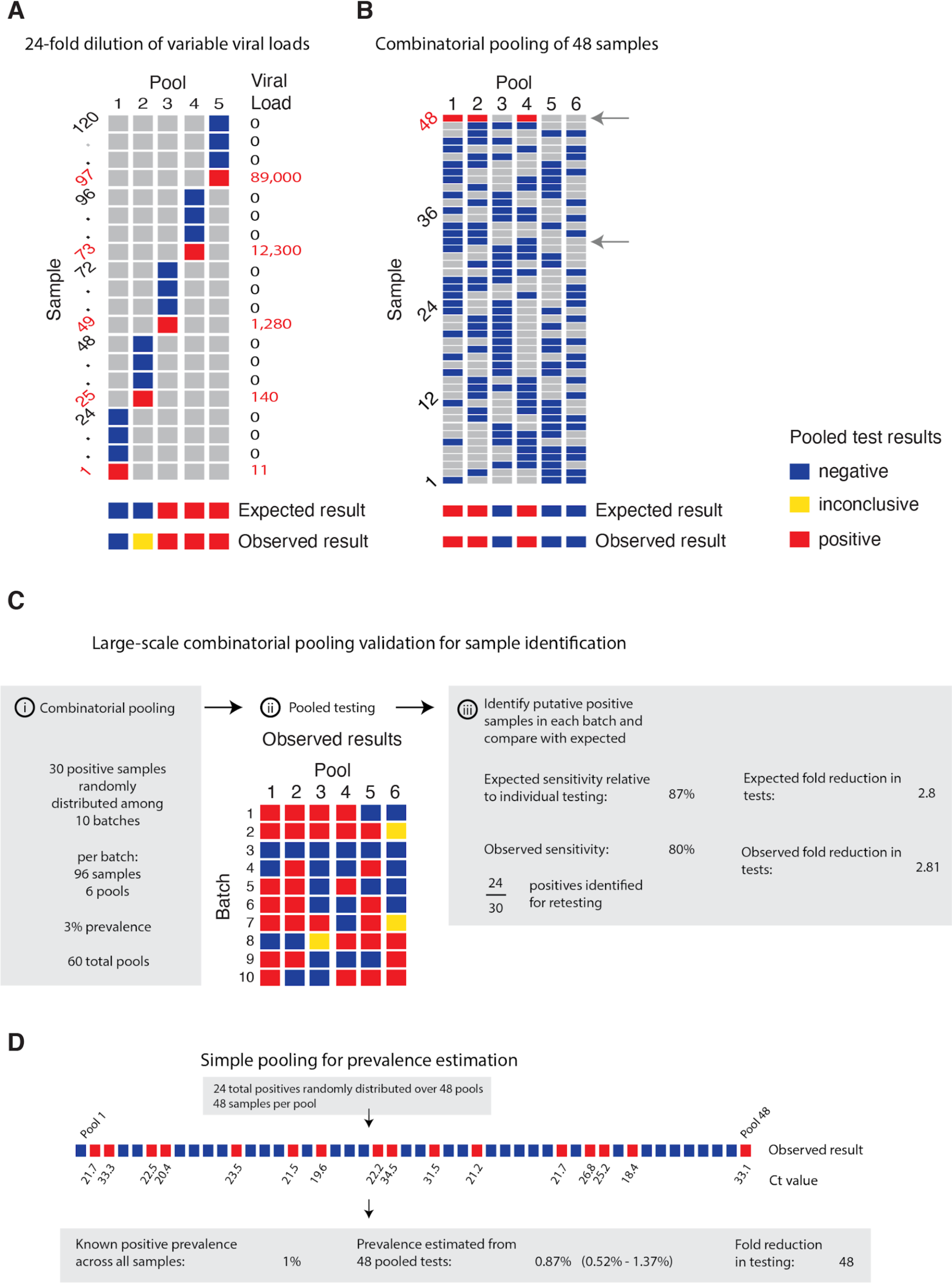
Experimental validation of simple and combinatorial pooling. (A) Five pools (columns of matrix), each consisting of 24 nasopharyngeal swab samples (rows of matrix; 23 negative samples per pool and 1 positive, with viral load indicated in red on right) were tested by viral extraction and RT-qPCR. Pooled results indicated as: negative (blue), inconclusive (yellow), or positive (red). (B) Six combinatorial pools (columns) of 48 samples (rows; 47 negative and 1 positive with viral load of 12,300) were tested as above. Pools 1, 2, and 4 tested positive. Arrows indicate two samples that were in pools 1, 2, and 4: sample 32 (negative), and sample 48 (positive). (C) Previously tested de-identified samples were pooled using a combinatorial design with 96 samples, 6 pools, and 2 pools per sample. 30 positive samples were randomly distributed across 10 batches of the design. Viral RNA extraction and RT-qPCR were performed on each pool, with the results used to identify potentially positive samples. (D) Samples were pooled according to a simple design (48 pools with 48 samples per pool). 24 positive samples were randomly distributed among the pools (establishing a 1% prevalence). The pooled test results were used with an MLE procedure to estimate prevalence (0.87%), and bootstrapping was used to estimate a 95% confidence interval (0.52% - 1.37%).

We next tested combinatorial pooling, first using only a modest pooling design. We split 48 samples, including 1 positive with a viral load of 12,300, into 6 pools with each sample spread across three different pools. The method correctly identified the three pools containing the positive specimen (Fig. 5B, **table S2**). One negative sample was included in the same 3 pools as the positive sample; thus, 8 total tests (6 pools + 2 validations) were needed to accurately identify the status of all 48 samples, a 6x efficiency gain, which matched our expectations from the simulations.

We next performed two larger validation studies (**Materials and Methods, Supplementary Material 1, section 10**). To validate combinatorial pooling, we used anonymized samples representing 930 negative and 30 distinct positive specimens (3.1% prevalence), split across 10 batches of 96 specimens each (**table S3**). For each batch of 96, we split the specimens into 6 pools and each specimen was spread across 2 pools (Fig. 5C; see **table S4** for sample allocation and **table S6** for pooled test results). For this combinatorial pooling design and prevalence, our simulations suggest that we would expect to identify ∼26 out of 30 known positives (∼87%) and would see a 2.81x efficiency gain – using only 35% of the number of tests compared to no pooling. We identified 24 of the 30 known positives (80%) and, indeed, required 35% fewer tests (341 vs 960, a 2.8x efficiency gain). Regarding the slightly reduced sensitivity, we note that positive samples and negative samples were accumulated over a longer period of time by the Broad’s testing facility, aggregated into separate plates, then delivered to our lab for pooling. Many of these samples went through multiple freeze-thaw cycles, likely leading to degradation of some viral RNA, which may have contributed to the reduced sensitivity that we observed relative to predicted levels.

To further validate our methods for prevalence estimation, we created a large study representing 2,304 samples with a (true) positive prevalence of 1%. We aimed to determine how well our methods would work to estimate the true prevalence using 1/48^th^ the number of tests compared to testing samples individually. To do this, we randomly assigned 24 distinct positive samples into 48 pools, with each pool containing 48 samples (**table S3**; to create the full set of pools, we treated some known negatives as distinct samples across separate pools). By using the measured viral loads detected in each of the pools, our methods estimated a prevalence of 0.87% (compared to the true prevalence of 1%) with a bootstrapped 95% confidence interval of 0.52% - 1.37% (Fig. 5D), and did so using 48x fewer tests than without pooling. This level of accuracy is in line with our expectations from our simulations. Notably, the inference algorithm applied to these data used viral load distributions calibrated from our simulated epidemic, which in turn had viral kinetics calibrated to samples collected and tested on another continent, demonstrating robustness of the training procedure.

## Discussion

Our results show that group testing for SARS-CoV-2 can be a highly effective tool to increase surveillance coverage and capacity when resources are constrained. For prevalence testing, we find that fewer than 40 tests can be used to accurately infer prevalences across four orders of magnitude, providing large savings on tests required. For individual identification, we determined an array of designs that optimize the rate at which infected individuals are identified under constraints on sample collection and daily test capacities. These results provide pooling designs that maximize the number of positive individuals identified on a daily basis, while accounting for epidemic dynamics, viral kinetics, viral loads measured from nasopharyngeal swabs or sputum, and practical considerations of laboratory capacity.

While our experiments suggest that pooling designs may be beneficial for SARS-CoV-2 surveillance and identification of individual specimens, there are substantial logistical challenges to implementing theoretically optimized pooling designs. Large-scale testing without the use of pooling already requires managing thousands of specimens per day, mostly in series. Pooling adds complexity because samples must be tracked across multiple pools and stored for potential re-testing. These complexities can be overcome with proper tracking software (including simple spreadsheets) and standard operating procedures in place before pooling begins. Such procedures can mitigate the risk of handling error or specimen mix-up.

In addition, expecting laboratories to regularly adapt their workflow and optimize pool sizes based on prevalence may not be feasible in some settings. (8, 15) A potential solution is to follow a simple, fixed protocol that is robust to a range of prevalences. We provide an example spreadsheet (**Supplementary Material 2**) guiding a technician receiving 96 labeled samples to create 6 pools, enter the result of each pool and be provided a list of putative positives to be retested.

For certain viscous sample types (e.g., saliva or sputum) the process of pooling samples may be challenging. While robotic pooling with small volumes of viscous samples is error-prone, manual pipetting can be reliable. Our designs can accommodate both robotic or manual pipetting, though pooling may still be a challenge for viscous samples compared to swabs in VTM or similar. In these cases, pre-treating samples with proteinase K can additionally make samples easier to pipette (as is done with SalivaDirect) and increase retention of viral RNA (by degrading nucleases). (23) Overall, our results are largely applicable to a variety of sampling landscapes in which viral loads are determined. However, individual laboratories will of course need to determine which approach is best suited for their own purposes.

Depending on the purpose of testing and resources available, enhancing efficiency at the expense of sensitivity must be considered. We recommend validation of the selected pooling strategy to identify potential differences in predicted versus observed sensitivity as demonstrated here, which may be unacceptable if the aim is clinical testing rather than overall throughput. However, we found that the most efficient pooling designs were the same when using swab- or sputum-calibrated simulations, suggesting that the ideal strategies were robust to misspecification in overall sensitivity. For prevalence testing, accurate estimates can be obtained using very few tests if individual identification is not the aim. For individual testing, although identifying all positive samples that are tested is of course the objective, increasing the number of specimens tested when sacrificing sensitivity may be a crucially important trade-off.

As an example of how to evaluate the allocation of limited resources in the context of reduced sensitivity and enhanced efficiency, consider our experience with a laboratory in The Gambia. Simplifying a bit, the resource constraints being faced were: (i) a limited capacity for sample collection (several hundred per day, initially – this has since increased); (ii) a limited supply of test kits for the foreseeable future; and (iii) an excess of laboratory technician capacity. For the purposes of screening, we assumed that up to 384 samples could be collected and up to 24 test kits consumed per day (on average, as there is a fixed supply over a longer horizon; sample collection and testing for triaging or testing higher-risk individuals was handled separately). With individual testing, effectively 20.4 individuals (24 available tests * 85% sensitivity) would be screened per day. On the other hand, with the pooling strategy identified under these constraints (Fig. 4c, N=192, b=12, q=3), one could effectively screen 202.57 individuals per day, on average (192 individuals screened per batch * 68% sensitivity * average of 1.55 batches run per day). Therefore, despite substantially reduced sensitivity, the pooling strategy will identify about 10x as many infected individuals. Given the unused technician capacity, the logistical cost (in our experience it takes one person ∼1.5 hours to pool 192 samples by hand in an unoptimized workflow, though there are other costs to consider) is very likely outweighed by the positive benefits on infection control.

Furthermore, the specimens most likely to be lost due to dilution are those samples with the lowest viral loads already near the limit of detection (**fig. S3D,E**). Although there is a chance that the low viral load samples missed are on the upswing of an infection – when identifying the individual would be maximally beneficial – the asymmetric course of viral titers over the full duration of positivity means that most false negatives would arise from failure to detect late-stage, low-titer individuals who are less likely to be infectious. (19) Optimal strategies and expectations of sensitivity should also be considered alongside the phase of the epidemic and how samples are collected, as this will dictate the distribution of sampled viral loads. For example, if individuals are under a regular testing regimen, and therefore likely to be removed from the tested population before reaching the long tail of low viral loads at the end of infection, or are tested due to recent exposure or symptom onset, then viral loads at the time of sampling will typically be higher, leading to higher sensitivity in spite of dilution effects.

For individual identification, errors may arise when a positive sample is split into multiple pools, but only some of those pools test positive (see **Supplementary Material 1, section 8** for an example and further discussion). For an error tolerance, *e*, error correction is possible by allowing a putative positive to be in up to *e* negative pools, where *e*>0 will result in higher sensitivity but also more putative positives and lower efficiency. The optimal error tolerance can be derived analytically in terms of the independent error rate r (*i.e.*, rates of PCR failure not related to low viral load, dilution or other systemic effects; **Materials and Methods**). (24) With a conservative estimate of the rate of independent PCR failure, *r*, of <5%, and samples split into less than 5 pools, *q*<5 (as in all pooling designs we consider), the optimal *e* is less than 0.5; thus, it is optimal to not correct these errors (e=0) in all pooling designs considered here. We note that for alternative pooling designs – particularly those with each sample split into many pools – error correction may increase effectiveness. Other methods that incorporate error correction (e.g., P-BEST (16)), have more complicated pooling designs with samples split into more pools (*q*=6 or more). An alternative to individually testing all putative positives would be to simply re-test each pool (optionally re-pooling samples before re-testing). Whether to do so or not would be a choice made by each lab, similar to decisions about whether to re-test samples with failed controls or inconclusive results. Furthermore, error correction does not address the primary source of error: low viral load, below the LOD and possibly due to dilution, which is a systemic problem affecting all pools with a given sample.

Testing throughput and staffing resources should also be considered. If a testing facility can only run a limited number of tests per day, it may be preferable to process more samples at a slight cost to sensitivity. Back-logs of individual testing can result in substantial delays in returning individual test results, which can ultimately defeat the purpose of identifying individuals for isolation - potentially further justifying some sensitivity losses. (25, 26) Choosing a pooling strategy will therefore depend on target population and availability of resources. For testing in the community or in existing sentinel surveillance populations (*e.g.,* antenatal clinics), point prevalence is likely to be low (<0.1 - 3%), which may favor strategies with fewer pools. (6,27–29) Conversely, secondary attack rates in contacts of index cases may vary from <1% to 17% depending on the setting (*e.g.*, casual vs. household contacts), (30–32) and may be even higher in some instances, (33, 34) favoring more pools. These high point prevalence sub-populations may represent less efficient use cases for pooled testing, as our results suggest that pooling for individual identification is inefficient once prevalence reaches 10%. However, group testing may still be useful if testing capacity is severely limited – for example, samples from all members of a household could be tested as a pool and quarantined if tested positive, enabling faster turnaround than testing individuals. This approach may be even more efficient if samples can be pooled at the point of collection, requiring no change to laboratory protocols.

Related to throughput, another important consideration is the overall time between sample collection and testing, particularly where individuals are advised to self-isolate until their result is returned. In settings where results are being returned within 24-48 hours already and there is no backlog of samples, pooling is unlikely to decrease turnaround time due to the extra time needed to retest some samples. However, in settings where testing capacity is insufficient to meet demand and results are delayed simply due to the queue of samples, pooling will likely decrease turnaround time because the time from sample to processing will be shorter.

Our modeling results have a number of limitations and may be updated as more data become available. First, our simulation results depend on the generalizability of the simulated Ct values, which were based on viral load data from symptomatic patients. Although some features of viral trajectories, such as viral waning, differ between symptomatic and asymptomatic individuals, population-wide data suggest that the range of Ct values do not differ based on symptom status. (19, 35) Furthermore, we have assumed a simple hinge function to describe viral kinetics. Different shapes for the viral kinetics trajectory may become apparent as more data become available. Nonetheless, our simulated population distribution of Ct values is comparable to existing data and we propagated substantial uncertainty in viral kinetics parameters to generate a wide range of viral trajectories. For prevalence estimation, the MLE framework requires training on a distribution of Ct values. Such data can be available based on past tests from a given laboratory, but care should be taken to use a distribution appropriate for the population under consideration. For example, training the virus kinetics model on data skewed towards lower viral loads (as would be observed during the tail end of an epidemic curve) may be inappropriate when the true viral load distribution is skewed higher (as might be the case during the growth phase of an epidemic curve). Nevertheless, we used our simulated distribution of Ct, which were fit to virus kinetics in published reports from distinct labs across the world and obtained highly accurate results throughout. Thus, despite the limitations just mentioned, this shows that virus kinetics models are quite robust and may not, in practice, require new fitting to the individual laboratory or population. In addition, while we assume that individuals are sampled from the population at random in our analysis, in practice samples that are processed together are also typically collected together, which may bias the distribution of positive samples among pools.

Pooling samples for qPCR testing is widespread in infectious disease surveillance, and our approach could be easily adapted to investigate pooled testing strategies *in silico* for other disease systems. (36–40) The viral kinetics model could be replaced with any model for the within-host process of interest (*e.g.,* an antibody response), and the epidemic model could be re-parameterized for particular settings (*e.g.,* a vector-borne with seasonal transmission). In general, modeling the key mechanisms behind the biological and epidemiological processes that give rise to measured quantities such as viral load provides a more nuanced picture of sensitivity and efficiency than treating test characteristics as fixed values.

We have shown that simple designs that are straightforward to implement have the potential to greatly improve testing throughput across the time course of the pandemic. These principles likely also hold for pooling of sera for antibody testing, which remains an avenue for future work. There are logistical challenges and additional costs associated with pooling that we do not consider deeply here, and it will therefore be up to laboratories and policy makers to decide where these designs are feasible. Substantial coordination will therefore be necessary to make group testing practical but investing in these efforts could enable community screening where it is currently infeasible and provide epidemiological insights that are urgently needed.

## Materials and Methods

### Simulation model of infection dynamics and viral load kinetics

We developed a population-level mathematical model of SARS-CoV-2 transmission that incorporates realistic within-host virus kinetics. Full details are provided in **Supplementary Material 1, sections 1-4,** but we provide an overview here. First, we fit a viral kinetics model to published longitudinally collected viral load data from nasopharyngeal swab and sputum samples using a Bayesian hierarchical model that captures the variation of peak viral loads, delays from infection to peak and virus decline rates across infected individuals (Fig. 2A**&B**; **fig. S10**). (18) By incorporating estimated biological variation in virus kinetics, this model allows random draws each representing distinct within-host virus trajectories. We then simulated infection prevalence during a SARS-CoV-2 outbreak using a deterministic Susceptible-Exposed-Infected-Removed (SEIR) model with parameters reflecting the natural history of SARS-CoV-2 (Fig. 2D). For each simulated infection, we generated longitudinal virus titers over time by drawing from the distribution of fitted virus kinetic curves, using distributions derived using either nasopharyngeal swab or sputum data (**fig. S4**). All estimated and assumed model parameters are shown in **table S5**, with model fits shown in **fig. S10.** Posterior estimates and Markov chain Monte Carlo trace plots are shown in **fig. S11** and **fig. S12.** We accounted for measurement variation by: i) transforming viral loads into Ct values under a range of Ct calibration curves, ii) simulating false positives with 1% probability, and, importantly, iii) simulating sampling variation. We assumed a limit of detection (LOD) of 100 RNA copies / ml.

### Estimating prevalence from pooled test results

We adapted a statistical (maximum likelihood) framework initially developed to estimate HIV prevalence with pooled antibody tests to estimate prevalence of SARS-CoV-2 using pooled samples. (41, 42) The framework accounts for the distribution of viral loads (and uncertainty around them) measured in pools containing a mixture of negative and potentially positive samples. By measuring viral loads from multiple such pools, it is possible to estimate the prevalence of positive samples without individual testing. See **Supplementary Material 1, section 5** for full details.

We evaluated prevalence estimation under a range of sample availabilities (*N total samples; N=*288 to ∼18,000) and pooling designs. We varied the pool size of combined specimens (*n samples per pool*; *n*=48, 96, 192, or 384) and the number of pools (*b*=6, 12, 24, or 48). For each combination, we estimated the point prevalence from pooled tests on random samples of individuals drawn during epidemic growth (days 20-120) and decline (days 155-300). Because the data is realistic but simulated, we used ground truth prevalence in the population and, separately, in the specific set of samples collected from the overall population to assess accuracy of our estimates (see for example Fig. 3B). We calculated estimates for 100 entirely distinct epidemic simulations.

### Pooled tests for individual sample identification

Using the same simulated population, we evaluated a range of simple and combinatorial pooling strategies for individual positive sample identification (**Supplementary Material 1, section 7**). In simple pooling designs, each sample is placed in one pool, and each pool consists of some pre-specified number of samples. If a pool tests positive, all samples that were placed in that pool are retested individually (Fig. 1A). For combinatorial pooling, each sample is split into multiple, partially overlapping pools (Fig. 1B). (9, 10) Every sample that was placed in any pool that tested negative is inferred to be negative, and the remaining samples are identified as potential positives. Here, we consider a very simple form of combinatorial testing, where identified potential positive samples are individually tested in a validation stage.

A given pooling design is defined by three parameters: the total number of individuals to be tested (*N*); the total number of pools to test (*b*); and the number of pools a given sample is included in (*q*). For instance, if we have 50 individuals (*N*) to test, we might split the 50 samples into four pools (*b*) of 25 samples each, where each sample is included in two pools. Note that, by definition, in simple pooling designs each sample is placed in one pool (*q*=1).

To identify optimal testing designs under different resource constraints, we systematically analyzed a large array of pooling designs under various sample and test kit availabilities. We evaluated different combinations of between 12 and ∼6,000 available samples/tests per day. The daily testing capacity shown is the daily average, though we assume that there is some flexibility to use fewer or more tests day to day (*i.e.,* there is a budget for period of time under evaluation).

For each set of resource constraints, we evaluated designs that split *N* samples between 1 to 96 distinct pools, and with samples included in *q*=1 (simple pooling), 2, 3, or 4 (combinatorial pooling) pools (**table S1**). To ensure robust estimates (especially at low prevalences of less than 1 in 10,000), we repeated each simulated pooling protocol at each time point in the epidemic up to 200,000 times.

In each scenario, we calculated: i) the sensitivity to detect positive samples when they existed in the pool; ii) the efficiency, defined as the total number of samples tested divided by the total number of tests used; iii) the total number of identified true positives (total recall); and iv) the effectiveness, defined as the total recall relative to individual testing.

### Pilot experiments

For validation experiments of our simulation-based approach, we used fully de-identified, discarded human nasopharyngeal specimens obtained from the Broad Institute of MIT and Harvard. In each experiment, sample aliquots were pooled before RNA extraction and qPCR and pooled specimens were tested using the Emergency Use Authorization (EUA) approved SARS-CoV-2 assay performed by the Broad Institute CLIA laboratory. qPCR cycle threshold values were calibrated to a standard curve of known viral RNA copies (**fig. S13**). The protocol is described in full detail in **Supplementary Material 1, section 9**. Specifics of each pooling approach is detailed alongside the relevant results and in **Supplementary Material 1, section 10**.

## Data Availability

Raw data generated in this study are available in Table S2 and Table S6. Simulated data can be regenerated using the accompanying code.

## Acknowledgements

We thank Benedicte Gnangnon, Rounak Dey, Xihong Lin, Edgar Dobriban, Rene Niehus and Heather Shakerchi for useful discussions. Work was supported by the Merkin Institute Fellowship at the Broad Institute of MIT and Harvard (B.C.), by the National Institute of General Medical Sciences (#U54GM088558; J.H. and M.M.); and by an NIH DP5 grant (M.M.), and the Dean’s Fund for Postdoctoral Research of the Wharton School and NSF BIGDATA grant IIS 1837992 (D.H.).

## Author contributions

B.C., J.H., A.R. and M.M. conceived the study. B.C. and J.H. performed the simulations and modeling. B.B., M.H, M.C, J.B, and B.S performed the pooled testing. D.H. and B.C. designed the combinatorial pools used for validation. M.S. and A.K.S. contributed to study design, discussion and analysis of effectiveness in a resource constrained environment. S.G. contributed to study design of validation experiments. B.C., J.H, A.R, and M.M. analyzed results and wrote the manuscript with input from all authors.

## Code availability

The SEIR model, viral kinetics model and MCMC were implemented in R version 3.6.2. The remainder of the work was performed in Python version 3.7. The code used for the MCMC framework is available at: https://github.com/jameshay218/lazymcmc. All other code and data used are available at: https://github.com/cleary-lab/covid19-group-tests.

## Data availability

Raw data generated in this study are available in **table S2** and **table S6**. Simulated data can be regenerated using the accompanying code.

## Supplementary Material 1: additional materials and methods

### 1. Model of infection dynamics

We implemented a deterministic compartmental model to describe the increase and subsequent decline of SARS-CoV-2 infection incidence and prevalence. The model captured the following progression: individuals begin fully susceptible to infection (S); become exposed but not yet infectious (E); become infectious (I); and are finally removed (R). Our aim was to capture changes in incidence over time (increasing incidence rate before the peak and declining incidence thereafter) and the resulting population-level distribution of viral loads, and we therefore did not model additional complexity such as deaths, pre-symptomatic transmission or age-stratified outcomes. Although incorporating these mechanisms may alter the simulated epidemic dynamics for a given set of transmission parameters, we do not expect them to impact inference from the pooled testing analyses.

The model was defined by the following set of ordinary differential equations (ODEs):

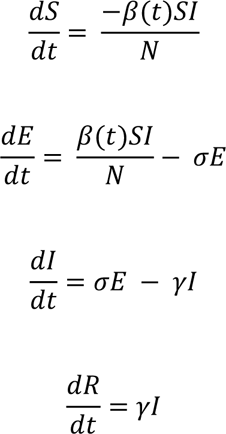

where *β*(*t*) is the time-varying transmission rate; 1/*σ* is the incubation period, assumed to have a mean of 6.4 days; 1/*γ* is the infectious period assumed to have a mean of 7 days; and *N* is the population size, set to 12,500,000 to generate at least 10,000,000 infected individuals. All model parameters and assumed values are described in **table S5**. Using this model, we simulated per capita daily infection probability (i.e., the daily incidence) and prevalence.

For the main analyses, we assumed that the transmission rate was held constant with *β*(*t*) = *R*_0_*γ*, where *R*_2_ is the basic reproductive number (assumed to be 2.5). For results in the section “Pooled testing in a sustained epidemic”, we assumed that the transmission rate changed over time as follows:

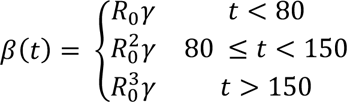

where *R*_2_ = 2.5, 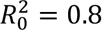 and 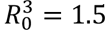. To account for gradual changes in transmission following the implementation or relaxation of interventions on days 80 and 150, we fitted a cubic smoothing spline to *β*(*t*) using the ‘smooth.spline’ function in R with a smoothing parameter of 0.5 before solving the ODEs.

### 2. Viral load kinetics

Following infection (*t*_inc_) and after a latent period (*t*_*g*_), viral load was assumed to increase exponentially, peaking a short time (*t*_*p*_) after the end of the latent period and before the onset of symptoms (*t*_*o*_). Viral load then decreased monotonically to undetectable levels, defined by a time parameter (*t*_*w*_) giving the number of days post symptom onset before crossing the limit of detection. These assumptions are equivalent to linear growth and decay on a log scale. We chose to model viral load waning with respect to symptom onset rather than peak viral load, as almost all available time-series viral load data are presented with respect to symptom onset time. The `log`_10_ viral load over time, *f*(*t*), was given by:

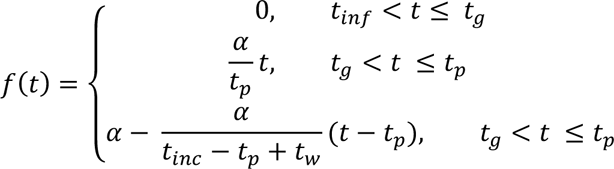

where *t*_inf_ is the time of infection; *t*_inc_ is the incubation period for symptom onset; *α* is the peak viral load; *t*_*g*_ is the latent period before viral growth; *t*_*p*_ is the time taken to reach peak viral load post; and *t*_*w*_ is the number of days from symptom onset to becoming undetectable. Note that all individuals are assigned a symptom onset time regardless of whether they show symptoms or not, as the parameter is used here to describe viral kinetics.

### 3. Fitting the viral kinetics model

Time-series viral load data were obtained from a case series of 9 hospitalized COVID-19 patients from a single hospital in Munich, Germany. (43) These data provide regular measurements of `log`_10_ RNA copies per nasopharyngeal swab and per ml of sputum. Data were extracted using a web plot digitizer (https://automeris.io/WebPlotDigitizer/). We developed a random-effects model to infer individual-level viral kinetics parameters alongside population-level distributions and fit this model separately to the swab and sputum data using a Markov chain Monte Carlo (MCMC) framework (**fig. S10**). All model parameters are summarized in **table S5**.

First, the observation level of the model is given by:

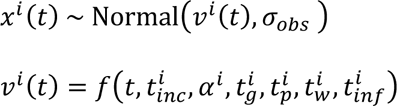

where *x*^*i*^(*t*) is the observed viral load for individual *i* at time *t* and *v*^*i*^(*t*) is the true viral load as defined above. Wölfel et al. present data with a limit of detection of 0 `log`_10_ copies / swab or / ml, but a limit of quantification of 2 `log`_10_ copies. We therefore define a likelihood function, *P*(*x*^*i*^(*t*)|*v*^*i*^(*t*)), assuming that the observed viral load is a normally distributed variable, *g*(*s*), with mean *v*^*i*^(*t*) and variance *σ*_*obs*_. The likelihood function accounts for censoring between the limit of detection and the limit of quantification:

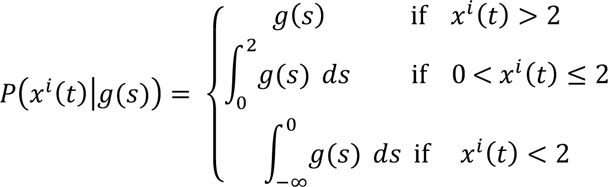

Next, we define the random-effects level on the individual-level viral kinetics parameters as:

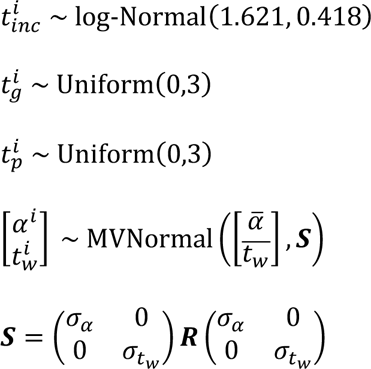

where MVNormal is the multivariate normal distribution. Finally, we define the remaining priors as:

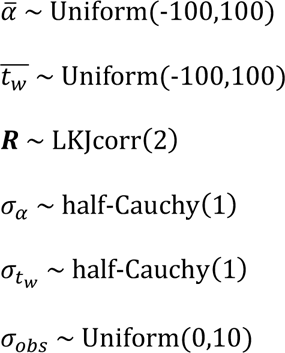

where 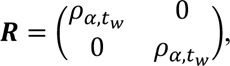, LKJcorr defines the LKJ correlation matrix distribution, and *t*_*in*f_ = 0 for all individuals.

Although the viral load measurements were taken after the onset of symptoms, we are interested in modeling the entire time course from the time of infection, including uncertainty in the incubation period. We therefore estimated *t*_inc_ for each individual by placing a strongly informative log-normal prior on the incubation period based on previous estimates. (44) Because no observations were made before symptom onset, the posterior distribution for this parameter matched the prior.

Code to regenerate the MCMC fitting is available in the accompanying GitHub repository. (45) Briefly, an adaptive Metropolis-Hastings algorithm was run for 3,000,000 iterations with the first 1,000,000 iterations discarded as burn in to obtain posterior estimates for the model parameters (**fig. S11**). Convergence was assessed visually based on trace plots of 3 independent chains (**fig. S12**). All estimated parameters had an effective sample size (ESS) greater than 200 and *R*^É^ < 1.1.

### 4. Simulation framework

We combined the estimates from the transmission model and viral kinetics model above to simulate viral loads for a population of 12,500,000 individuals over time. We performed separate simulations for swab and sputum data. For each individual, we simulated a time of infection (or no infection), *t*_*inf*_ from the infection incidence curve, *p*(*t*). We then drew viral kinetics parameters and an incubation period,*t*_*inc*_, for each individual from the estimated joint posterior distribution of the model parameters as defined above. Finally, we simulated the time-varying viral load of each infected individual, starting at the time of infection. Because of the high variance of the drawn kinetics parameters, some simulated viral loads reached very high values, and we therefore truncated all simulated viral loads at 16 `log`_10_ virus copies per ml (only 0.0025% of simulated swab viral loads are above this threshold, suggesting their impact on our results is very minor). These simulations provide a population of viral loads incorporating realistic variability in viral load kinetics and between-individual parameters.

### 5. Framework for prevalence estimation from pooled test results

In this section, we adapt a previously described maximum likelihood estimate (MLE) framework to estimate prevalence of positive samples, *p*, amongst *N* samples using *b* pooled tests, with each pooling containing 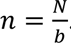. (41, 42) Briefly, the MLE is determined by computing the conditional probability of an observed RT-qPCR result given that there were *k* positive samples in the pool, and integrating over all values of *k*. To compute these conditional distributions, we estimate an empirical distribution of cycle threshold (Ct) values that follows from the distribution of viral loads in infected individuals derived from a subset of the simulated viral load measurements described above. This is conceptually similar to using the empirical distribution of all real measurements from a population taken to date.

Let *f*_*k,n*_ = *P*(*y*_i_|*k,n*) represent the conditional probability of the observed Ct values in each pool, *Y* = [*y*_1_, …, *y*_*b*_], given that there were *k* positive samples in the pool and *n-k* negative samples. Computing these distributions will be the central task of generating prevalence estimates.

To begin, we model Ct values as a function of the underlying viral loads:

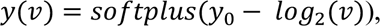

where *v* is the viral load (in copies per ml), and *softplus(Ct)* = *log*(1 + *e Ct*) is applied so that small Ct values smoothly decay toward zero, while larger Ct values still obey a linear relationship with log viral load (without this transformation, very large viral loads would imply nonsensical, negative Ct values). To capture variance in the LOD between labs or across batches within a lab, we let the intercept, *y*_0_, be normally distributed with unknown mean and standard deviation. We assume uniform priors over each *y*_0_ μ ~ *uniform*(37,39), σ ~ *uniform*(0.1,1)). These ranges and the log-linear relationship are based on a calibration of Ct values to a dilution curve of positive control SARS-CoV-2 RNA samples (**fig. S13**) and prior measurements calibrated to SARS coronavirus. (46)

We approximate *f*_*k,n*_ through a kernel density estimate (KDE) of empirical convolutions, while assuming that viral loads in uninfected individuals are always zero, and allowing for false positives, as discussed below. For a given value of *n* and each value of *k* from 1 to *n*, we generate Ct values *X*_*k,n*_ from 10,000 random combinations of *k* viral loads as follows:

*For i from* 1 *to* 10,000:

i. Sample net viral load 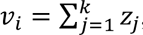, where *z*_*j*_ are viral loads sampled uniformly from all available training data
ii. Sample a Ct intercept value from μ ~ *uniform*(37,39), σ ~ *uniform*(0.1,1) and *y*_0_ ~ *N*(μ,σ)
iii. Convert viral load to Ct value 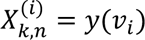

We then approximate *f*_*k,n*_ = `KDE`(*X*_*k,n*_) with a bandwidth parameter of 0.1.

Several adjustments to these density approximations are needed to account for false positives and undefined Ct values (*i.e.*, undetected viral RNA). To allow for Ct values that might arise from false positives, we define *f*_0,n_ = *f*_1,n_ *r*, where *r* is an assumed false positive rate of PCR (here, we set *r* = 0.2% and allow this to be misspecified when simulating false positives below). When viral RNA is undetected and *k* > 1, we let 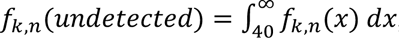, where a Ct value of 40 is used as a typical limit of detectable RNA. When *k* = 0, *f*_0,n_(`undetected`) = 1 − *r*.

Following the model of Zenios and Wein, (41) the MLE of prevalence, *p*, is defined as:

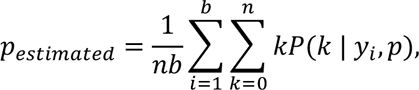

where *n* is the number of samples per pool, *b*, is the number of pools, and *y*_>_ is the Ct value observed in pool *i*. We calculate *P*(*k* | *y*_>_, *p*) using the conditional densities above:

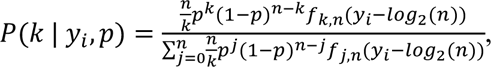

substituting the appropriate values of *f* when the Ct value is undefined. Note that each of the observed Ct values is adjusted to account for *n*-fold dilution by subtracting `log`_2_(*n*) from each. Finally, we find *p*_estimated_ through an iterative expectation-maximization algorithm, where at each iteration

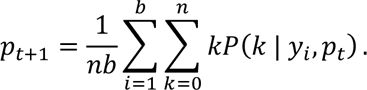

### 6. Prevalence estimation simulations

Using this model, we evaluated prevalence estimation across time in our simulated populations. We first partition the simulated population into training and testing sets. Observed viral loads in 20% of individuals (selected at random) are used to train the KDEs above. Furthermore, training data were divided into growth phase (days 20 to 120) or decline phase (days 155 to 300) data, allowing us to ascertain the effect of training and estimating on data from consistent or inconsistent phases of the epidemic. All remaining prevalence estimation and analyses are done on the testing population.

We simulate viral loads in *b* pools of samples as 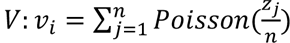 for *i* from *1* to *b*, where *z* are randomly sampled viral loads and Poisson sampling is used to model the sampling of a small volume from each swab. From these we simulate Ct values, *Y*. At each time point, we randomly select *N* individuals, partition them into *b* pools, and simulate the pooled viral loads and Ct values, *Y*, with *y*_2_ ∼ *N*(38.5,1) sampled randomly in each trial and applying the transformation described previously. To capture false positive PCR results at a rate of 1%, with 1% probability in each pool we add a viral load of 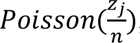, with *z* selected at random from all positive samples with 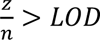 on a given date.

We then compute the true prevalence in the population, *p*_true_, the prevalence in the population of *N* samples, *p*_sampled_, and the prevalence estimated from *b* tests, *p*_estimated_. To quantify accuracy we calculate the mean and standard deviation of these values at each time point across 100 random trials and summarize differences between *p*_estimated_ and either *p*_true_ or *p*_sampled_ with the normalized rootmean squared error (NRMSE; either 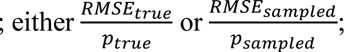; **fig. S2**).

To demonstrate the importance of calibrating the correct empirical distribution of Ct values, **fig. S2** shows the accuracy of the framework in recovering the true population prevalence during the growth and decline phases of the epidemic when the model is calibrated to Ct values measured during the growth and decline phases. Estimation accuracy is highest when the calibrated Ct values were measured during the same phase as is being measured.

### 7. Group testing for sample identification

We evaluated an array of pooling designs for individual identification as described in the main text. For any given design we can simulate pooled testing results on the simulated population described above.

For each time point, we randomly select *N* individuals from the population. Each individual is then randomly assigned to *q* out of *b* pools. For each pool, we then calculate the net sampled viral load, *v*_*i*_ = 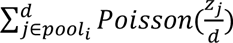 using the viral loads *z* for each individual included in `pool`_i_, where Poisson sampling is used to model a small volume sampled from each swab. If *v*_i_ > `LOD` the pool is assigned a positive value, otherwise the pool is negative (here, with *LOD*=100). To allow for false positive PCR results at a rate of 1%, with probability 1% we set the result of the pool to be positive, regardless of the viral load.

Simulations were run by repeating the random pooling, pooled testing, and decoding procedures described in the main text. At each point in time, 200,000 trials are run selecting *N* individuals at random in each trial. We then record the number of validation tests, *s*, in each trial, corresponding to the number of putative positive samples. Average efficiency at a point in time, *t*, is then calculated as 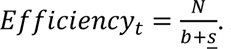. Average sensitivity is calculated as 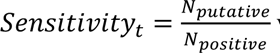 is the total number of infected individuals (viral load > 1) that were sampled across all trials and *N*_positive_ is the number of such individuals who were identified as putative positives. From these values we calculate Total Recall_t_ = Efficiency_t_ Sensitivity_t_.

Based on these results we evaluated a large array of group testing designs (**table S1**) under a series of constraints on number of samples collected and number of reactions run per day (Fig. 4C). For a given design, *D*: (*N, b, q*) we calculate the average number of tests run, *s*_total_ = *b* + <u>*s*</u>, and the total number of times that *D* could be run on a daily basis, 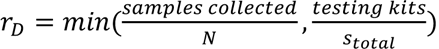. We then calculate the effective testing capacity on a daily basis, *c*_*D*_ = *N r*_*D*_ `Sensitivity`_*D*_, and average this value over days 40 to 90 (Fig. 4C) or 80 to 108 (**fig. S6**) of the simulated epidemic. Notably, when sample collection is limiting, *c*_*D*_ = samples collected * Sensitivity_(samples collected,, samples collected, 1)_, corresponding to individual testing. Finally, the optimal design for a given combination of constraints is the design with the greatest average effective testing capacity. In Fig. 4C and fig**. S6** we show the relative effectiveness, corresponding to effective capacity relative to individual testing. **fig. S7** shows the same results as Fig. 4 and fig**. S6**, but when samples were simulated based on viral trajectories using the sputum data. **fig. S5** shows the same results again, but for the decline phase of the epidemic.

### 8. Note on error correction for sample identification

Error correction is possible in our framework by allowing each putative positive to be in up to *e* negative pools. All results we present used *e* = 0 while technically allowing for the possibility of error correction (i.e., *e* > 1). Below we explain how this choice was made, based on the likely low rate of independent PCR failure, r.

Our simulations allowed for the possibility of error correction by following a theoretical formula on the optimal number of errors to tolerate; specifically, *e* ≈ 2*rq*. (24) With r<5% (a conservative estimate of the rate of independent PCR failure) and q<5 (as in all pooling designs we consider) the optimal e is less than 0.5, which we round down to 0; thus, it is optimal to not perform additional confirmatory tests to account for these errors.

The following example compares the results of e=0 with e=1 for the particular pooling design used in our large-scale validation experiments (with n=96, m=6, q=2; **table S3**). For simplicity, we assume a single positive test and a single positive sample, which, without error, would have produced two positive tests (because every sample is in q=2 pools). Without error correction, there are no putative positive samples. With e=1, every sample in the pool that tested positive is a putative positive. Each pool in this design has 32 samples; therefore, no matter which pool is positive, the cost of identifying a single sample that would have been missed without error correction is 32 additional tests (since putative positives are individually validated in stage ii). The marginal gain in sensitivity is greatly outweighed by the loss in efficiency, and thus, overall effectiveness is greatly reduced in this instance, when applying error correction. We reiterate that for alternative pooling designs – particularly those with each sample split into many pools – error correction may increase effectiveness, but for all designs in our analysis this is not the case.

### 9. Broad Institute CRSP SARS-CoV-2 Real-time Reverse Transcriptase (RT)-PCR Diagnostic Assay

RNA is isolated from nasopharyngeal and oropharygneal specimens in 50ul VTM/UTM using MagMAX™-96 Viral RNA Isolation Kits (Thermo Fisher Scientific, AMB18365), performed on an Agilent Bravo Liquid Handling Platform running VWorks (Build 11.4.0.1233), and is reverse transcribed to cDNA and subsequently amplified in the Applied Biosystems® ViiA7 Real-Time PCR Instrument with QuantStudio version 1.3 software. In the process, the probe anneals to a specific target sequence located between the forward and reverse primers (using standard probes for N1, N2, and RP). During the extension phase of the PCR cycle, the 5’ nuclease activity of Taq polymerase degrades the probe, causing the reporter dye to separate from the quencher dye, generating a fluorescent signal. With each cycle, additional reporter dye molecules are cleaved from their respective probes, increasing the fluorescence intensity. Fluorescence intensity is monitored at each PCR cycle by Applied Biosystems® ViiA7 Real-Time PCR System with QuantStudio version 1.3 software. The software allows the fluorescence intensity to be monitored at each PCR cycle to allow for the qualitative detection of nucleic acid from SARS-CoV-2. Using this protocol a calibration curve connecting viral copies to Ct value was generated through a dilution series of synthetic RNA (Twist BioScience Covid synthetic RNA) (**fig. S13**).

To create the final pools for our larger validation studies (10 batches of 96 for sample identification, and 48 pools of 48 samples for prevalence estimation), we treated a total of 96 known negative specimens as distinct samples across batches (in the case of sampled identification study) or pools (for the prevalence study). Each positive sample in a batch or pool was a distinct sample, and only used in one batch or pool. For the prevalence study, only pools with one or more positive samples were tested using the assay above (the remaining pools were assigned an “undefined” Ct value). All pools in all batches were tested in the identification study, regardless of whether they contained a positive sample.

### 10. Selecting pool compositions for large-scale sample identification validation

To form pools, we put each of the N = 96 individuals into *q* = 2 out of *b* = 6 pools (A-F) by cycling through the following ordered list of pool pairs: AB, CD, EF, BC, DF, AE, BD, AF, CE, BE, CF, AD, BF, DE, AC. Namely, individual 1 is put in pools A and B, individual 2 in C and D, and so on; after individual 15, we return to the beginning of the list and cycle through again. Finally, since each pair of pools is assigned to multiple individuals, we reordered the individuals by what pair of pools they were put in. This final reordering simplifies the presentation to one more conveniently arranged for manual pipetting.

A strength of the procedure is that it is simple and flexible; it can easily be carried out for any number of individuals. Moreover, the resulting design here has pools with 32 individuals each, so the final pool sizes are balanced. In addition, the first 6 pool pairs are each assigned to 7 individuals and the remaining 9 pairs are each assigned to 6 individuals, so the pairs of pools are also approximately balanced. Indeed, the ordered list of pool pairs above was chosen to achieve this balance; the list uses each pair of pools once and the five consecutive sets of three pairs all use each pool once.

---

**Fig. S1.**
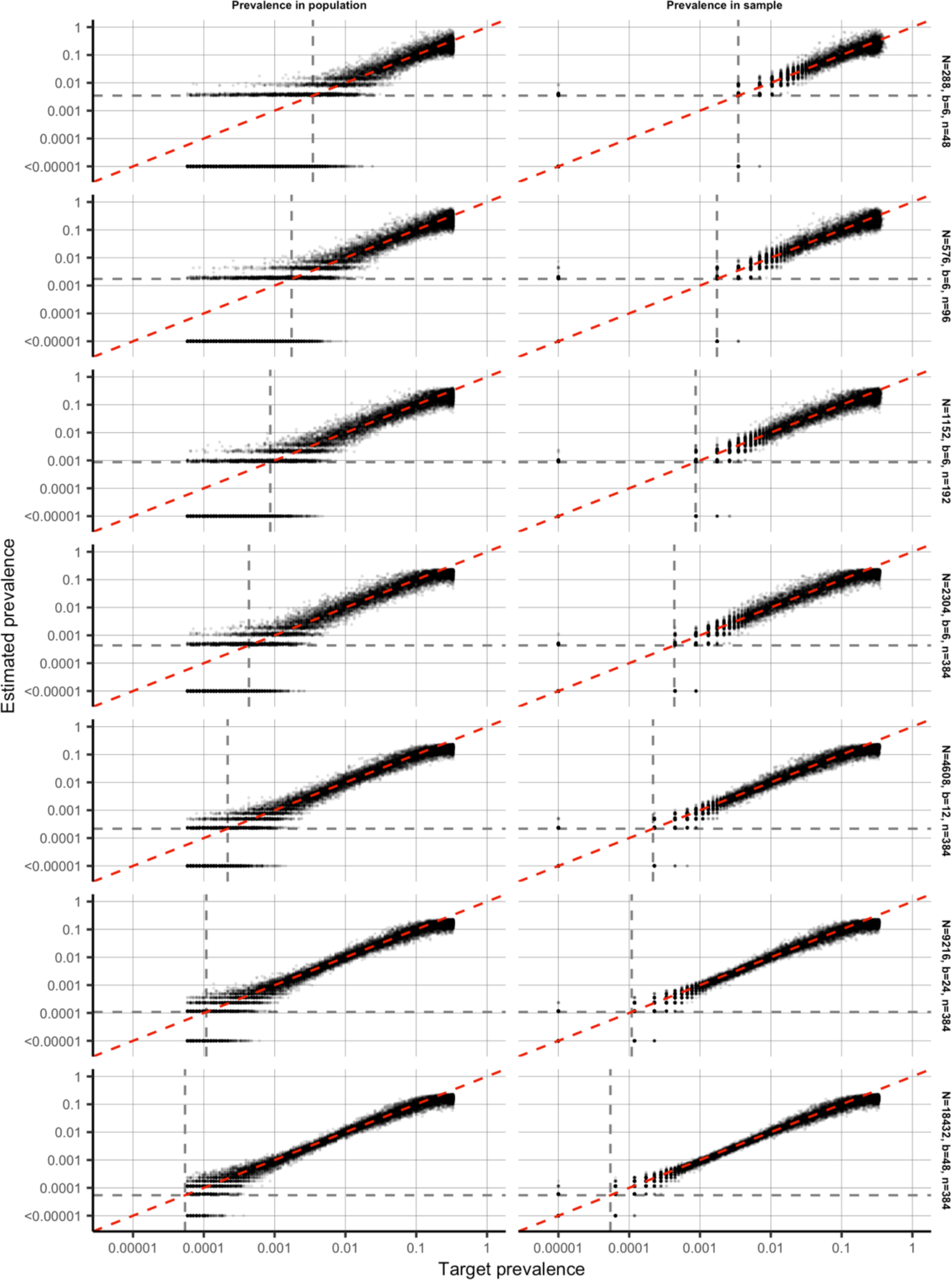
Population prevalence (left) or prevalence in sample (right) against maximum likelihood prevalence estimates. Population prevalence shown here are during the epidemic growth phase. Results shown are from 100 independent trials at each day of the epidemic. Each facet shows the pooling design with the fewest pools (tests used) for each sample size. Dashed grey lines show one divided by the sample size, N.

**Fig. S2.**
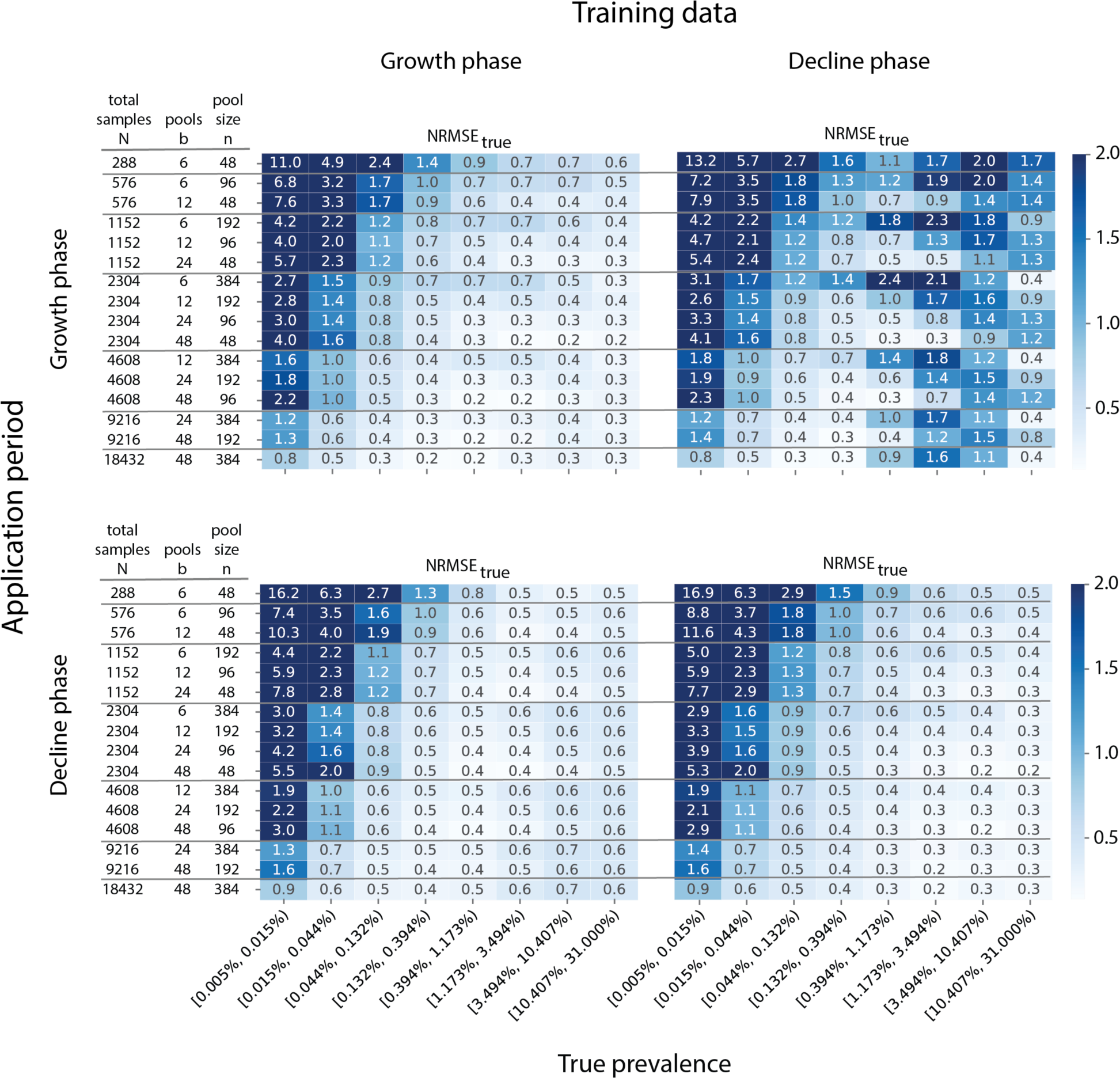
Prevalence estimation can depend on training and application period. Prevalence estimation uses past observations to learn conditional distributions of pooled Ct values. When the inference algorithm is applied to Ct values from new pooling data, these training distributions are used to generate a maximum likelihood estimate. The error in these estimates (here, NRMSE, the root mean squared error normalized to the true prevalence) depends on the pooling design (individual rows) and true prevalence (columns, binned prevalence windows). In addition, error can depend on a (mis)match between the training (two columns of panels) and application (two rows of panels) period. Colorbar and annotated values within boxes indicate NRMSE.

**Fig. S3.**
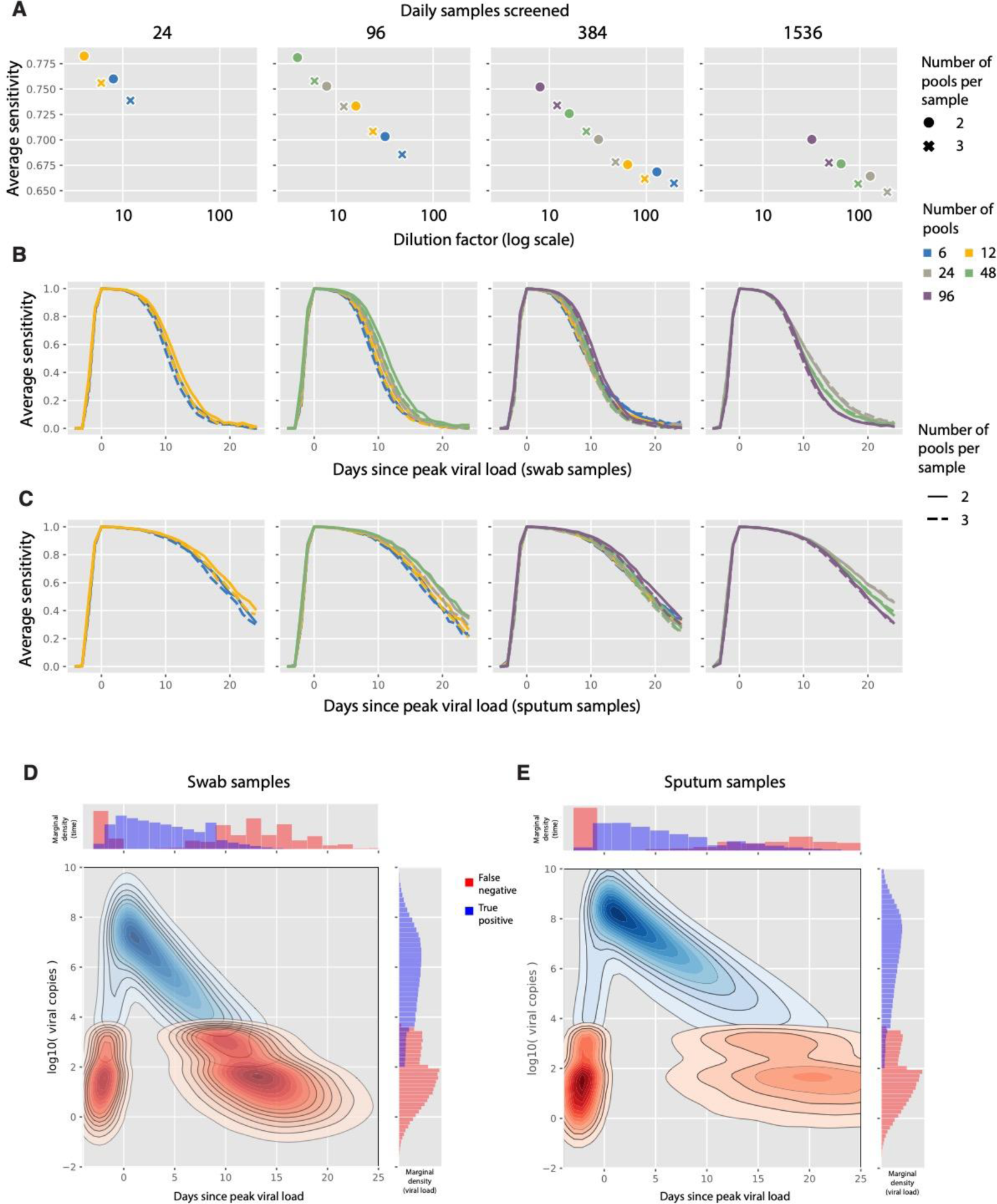
Sensitivity of sample identification relative to dilution factor and time since peak viral load. (A) The sensitivity (y-axis) of different designs (individual points) is plotted against the dilution factor of each design (x-axis, log scale). Pooling designs are separated by the number of swab samples tested on a daily basis (individual panels); the number of pools (color); and the number of pools into which each sample is split (circle versus cross). (B and C) As in (A), with sensitivity plotted against days since peak viral load (x-axis) for swab (B) and sputum (C) samples. (D and E) The density of false negative (red) and true positive (blue) results is shown as a function of days since peak viral load (x-axis) and viral load (y-axis, *log10* scale) at the time of sample collection and pooled testing for swab (D) and sputum (E) samples. Contour plots depict 2-d density, and histograms show marginal density over time (top) and viral load (right).

**Fig. S4.**
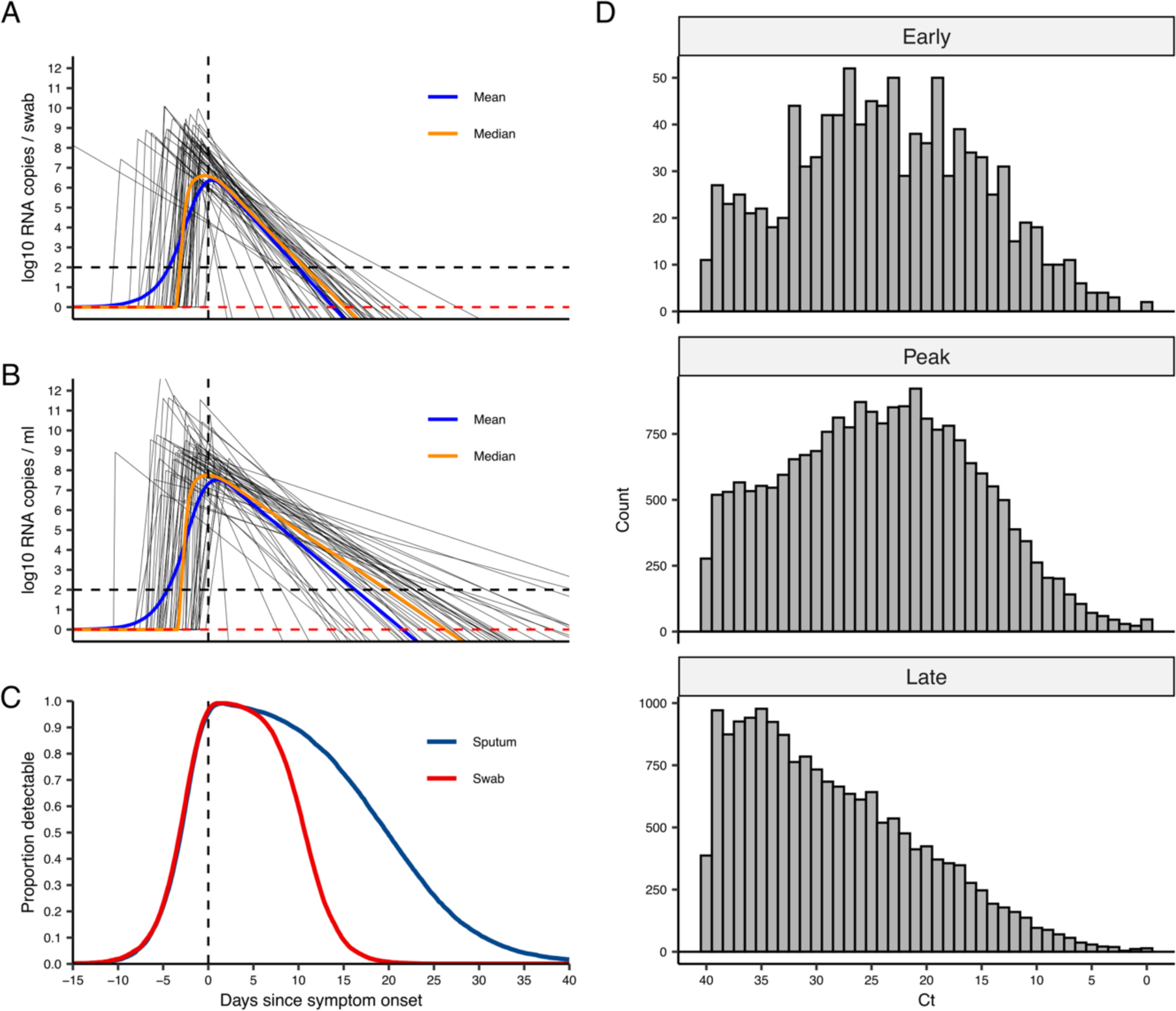
Simulated viral loads. (A) Black lines show 50 randomly drawn viral load trajectories based on parameters estimated from fitting to swab data. Vertical dashed line shows time of symptom onset. Horizontal dashed lines show limit of detection assumed in subsequent analyses (black line) and limit of detection reported by Wölfel et al (red line). Blue and orange line shows median viral load on each day with respect to symptom onset. (B) as in (A), but fitted to sputum data. (C) Proportion of true viral loads above the limit of detection (LOD = 2 *log10* RNA copies / swab or ml) on each day with respect to symptom onset for simulated swab (red) and sputum (blue) data. Note that the distribution for observed viral loads will differ slightly due to the addition of observation and sampling error. (D) Distribution of simulated cycle threshold values from swab samples at different stages of the epidemic.

**Fig. S5.**
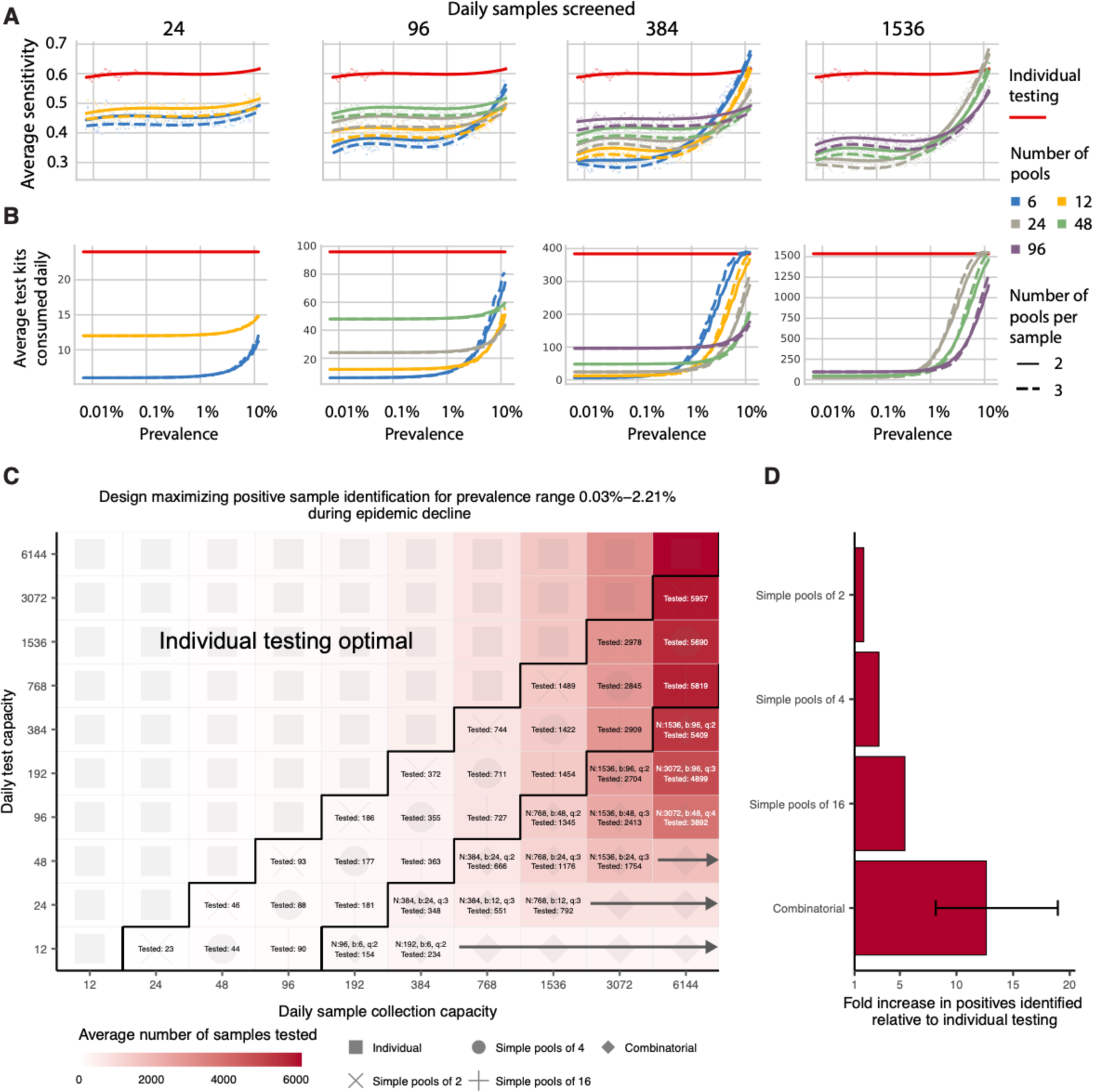
Group testing for sample identification during epidemic decline. Results shown are based on the same testing strategies as in Fig. 4. We evaluated a variety of group testing designs for sample identification (table S1) on the basis of sensitivity (A) shows sensitivity and (B) shows efficiency. total number of positive samples identified (C) and the fold increase in positive samples identified relative to individual testing (D). (A and B) The average sensitivity (A, y-axis, individual points and spline) and average number of tests needed to identify individual positive samples (B, y-axis) using different pooling designs (individual lines) were measured over days 20-110 in our simulated population, with results plotted against prevalence (x-axis, log-scale). Results show the average of 200,000 trials, with individuals selected at random on each day in each trial. Pooling designs are separated by the number of samples tested on a daily basis (individual panels); the number of pools (color); and the number of pools into which each sample is split (dashed versus solid line). Solid red line indicates results for individual testing. (C) Every design was evaluated under constraints on the maximum number of samples collected (columns) and average number of reactions that can be run on a daily basis (rows) over days 40-90. Text in each box indicates the optimal design for a given set of constraints (number of samples per batch (N), number of pools (b), number of pools into which each sample is split (q), average number of total samples screened per day). Color indicates the average number of samples screened on a daily basis using the optimal design. Arrows indicate that the same pooling design is optimal at higher sample collection capacities due to testing constraints. (D) Fold increase in the number of positive samples identified relative to individual testing with the same resource constraints. Error bar shows range amongst optimal designs.

**Fig. S6.**
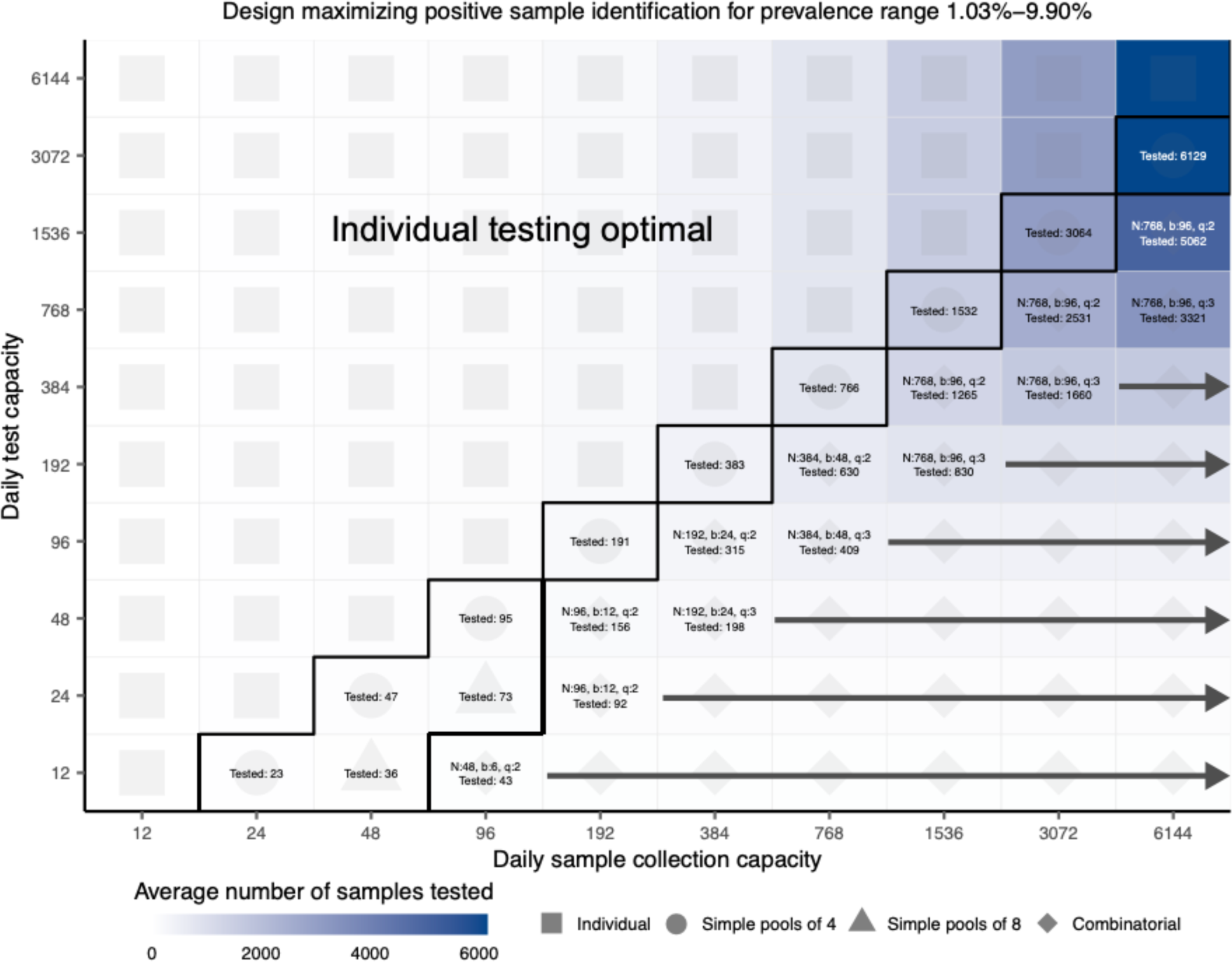
Effectiveness of optimal testing design under resource constraints at high prevalence. As in Fig. 4, every design was evaluated under constraints on the maximum number of samples collected (columns) and average number of reactions that can be run on a daily basis (rows), here from days 80-108. Text in each box indicates the optimal design for a given set of constraints (number of samples per batch (N), number of pools (b), number of pools into which each sample is split (n), average number of total samples screened per day). Color indicates the average number of samples screened on a daily basis using the optimal design. Arrows indicate that the same pooling design is optimal at higher sample collection capacities.

**Fig. S7.**
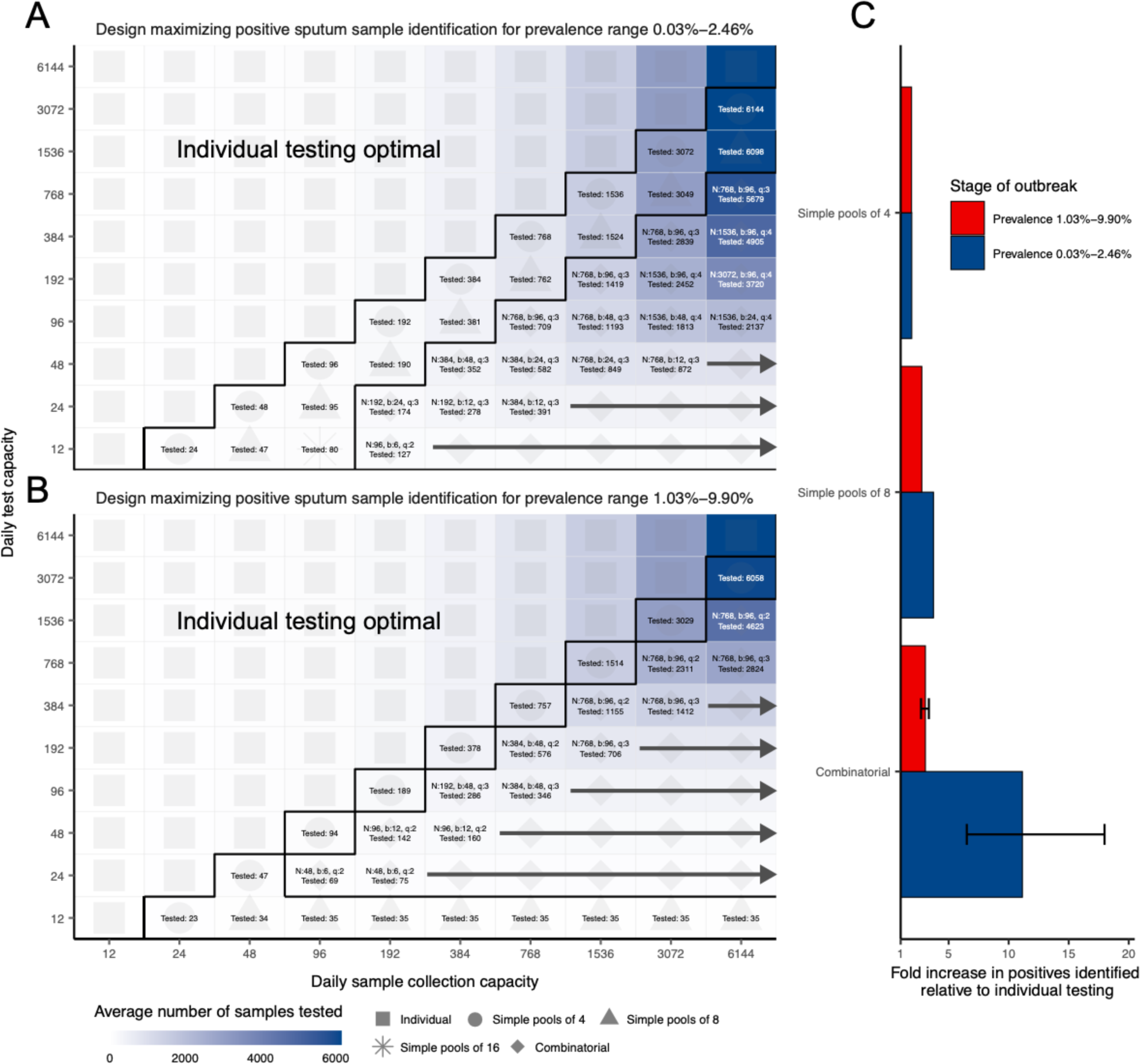
Effectiveness of optimal testing design under resource constraints using sputum data. (A) and (B) show optimal designs at lower and higher prevalence ranges respectively. Text in each box indicates the optimal design for a given set of constraints (number of samples per batch (N), number of pools (b), number of pools into which each sample is split (n), average number of total samples screened per day). Color indicates the average number of samples screened on a daily basis using the optimal design. Arrows indicate that the same pooling design is optimal at higher sample collection capacities. (C) Fold increase in the number of positive samples identified relative to individual testing with the same resource constraints. Error bar shows range amongst optimal designs.

**Fig. S8.**
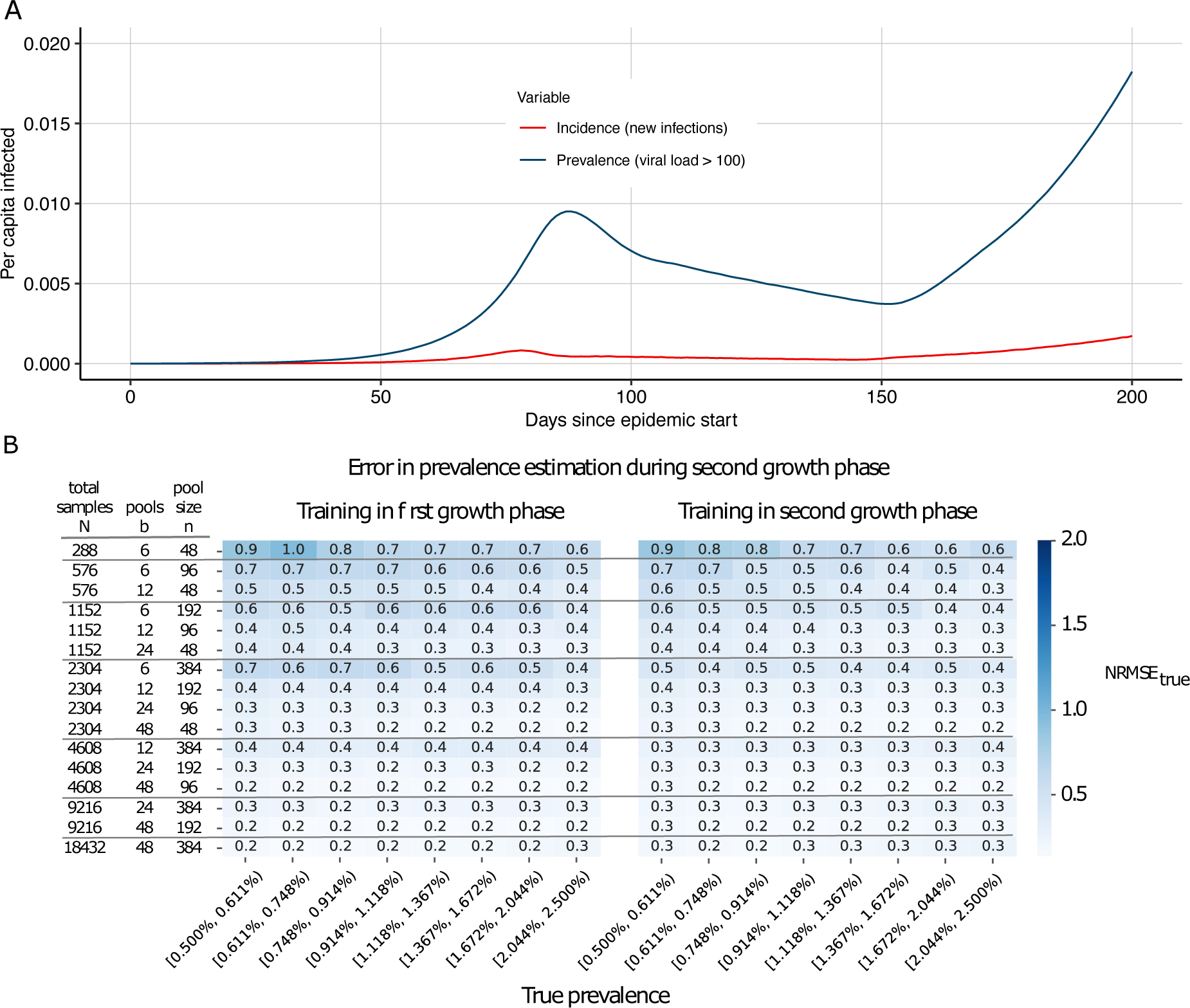
Evaluation of pooled testing in a sustained, multi-wave epidemic. (A) Simulated infection incidence and prevalence of virologically positive individuals from the SEIR model with two waves. Incidence (red line) was defined as the number of new infections per day divided by the population size. Prevalence (blue line) was defined as the number of individuals with viral load > 100 (log10 viral load > 2) in the population divided by the population size on a given day. (B) Error in prevalence estimation during the second growth phase using viral load distributions from training data in the first growth phase (up to day 80; left) or the second growth phase (day 150 onward; right). The error in these estimates (here, NRMSE, the root mean squared error normalized to the true prevalence) depends on the pooling design (individual rows) and true prevalence (columns, binned prevalence windows). Colorbar and annotated values within boxes indicate NRMSE.

**Fig. S9.**
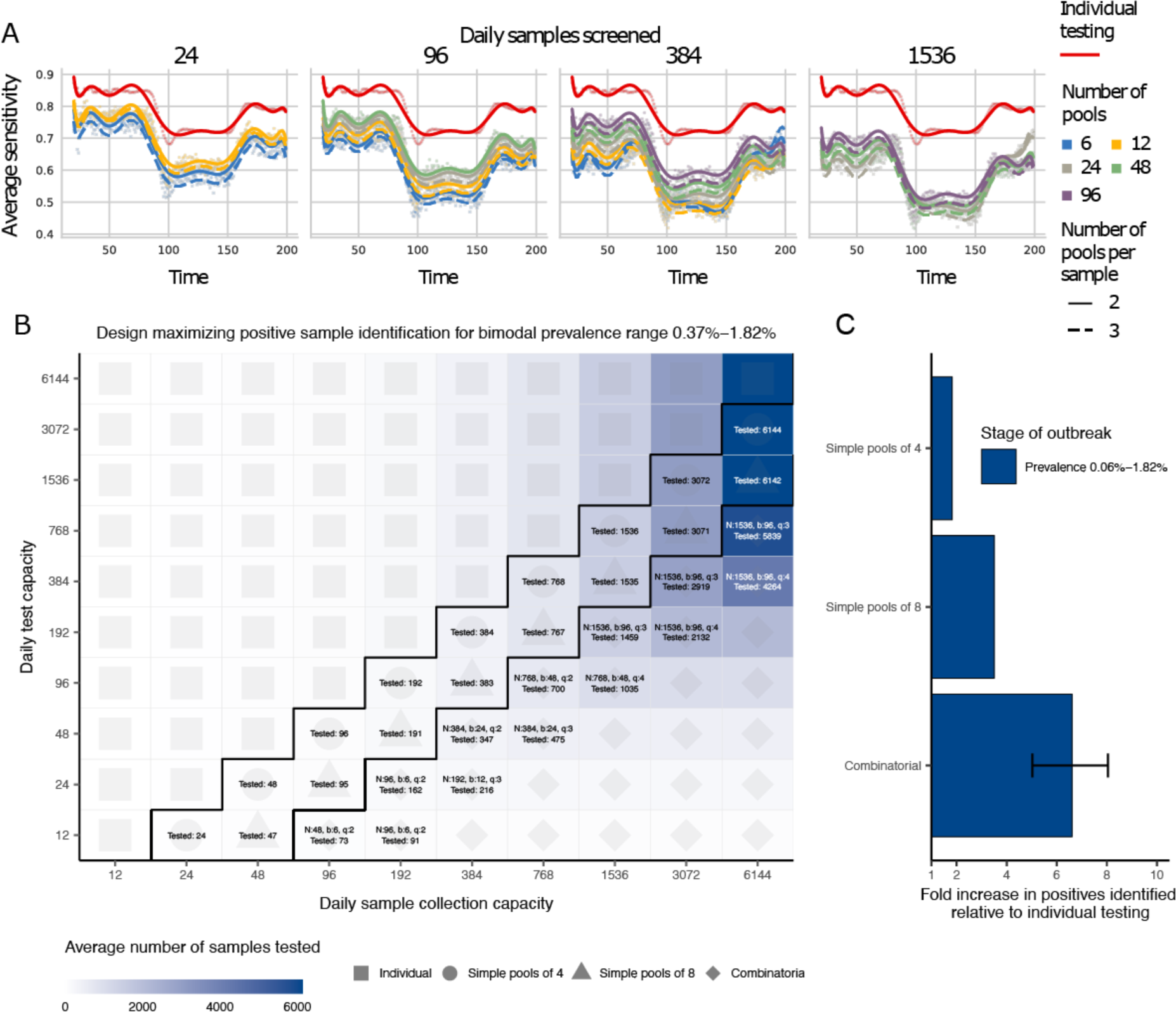
Evaluation of pooled testing for sample identification in the multi-wave epidemic shown in fig. S8a. (A) The average sensitivity (y-axis, individual points and spline) using different pooling designs (individual lines) were measured over days 20-200 (x-axis) in our simulated population. Results show the average of 200,000 trials, with individuals selected at random on each day in each trial. Pooling designs are separated by the number of samples tested on a daily basis (individual panels); the number of pools (color); and the number of pools into which each sample is split (dashed versus solid line). Solid red line indicates results for individual testing. (B) Every design was evaluated under constraints on the maximum number of samples collected (columns) and average number of reactions that can be run on a daily basis (rows) over days 150-200. Text in each box indicates the optimal design for a given set of constraints (number of samples per batch (N), number of pools (b), number of pools into which each sample is split (q), average number of total samples screened per day). Color indicates the average number of samples screened on a daily basis using the optimal design. (C) Fold increase in the number of positive samples identified relative to individual testing with the same resource constraints. Error bar shows range amongst optimal designs.

**Fig. S10.**
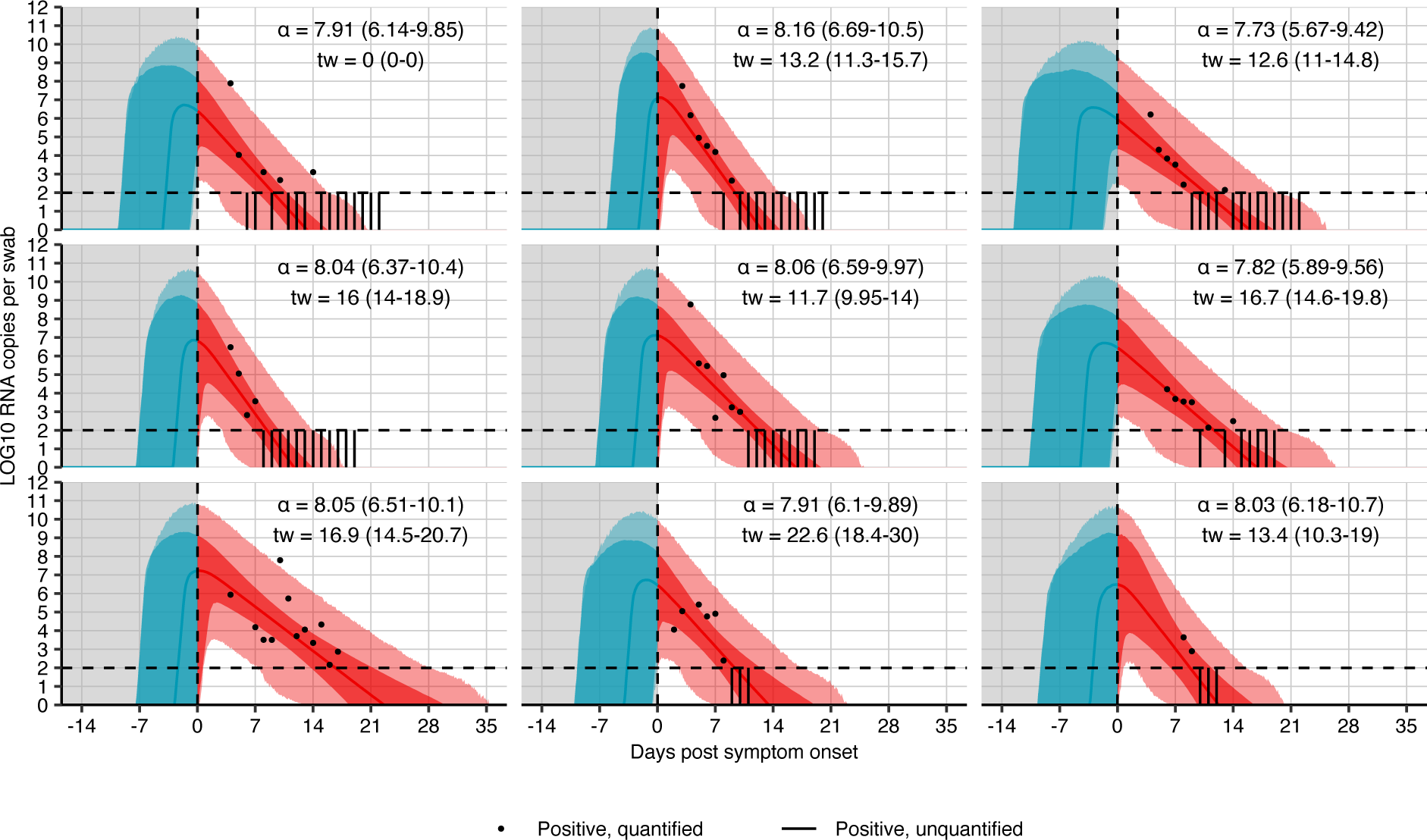
Model fits to swab viral loads. Data were extracted from Wölfel et al. 2020. (43) The black dots show observed *log10* RNA copies/swab; the vertical bars show positive, unquantified swabs; solid lines show posterior median estimates; dark shaded regions show 95% credible intervals (CI) on model-predicted latent viral loads; light shaded regions show 95% CI on simulated viral loads with added observation noise. The vertical dashed line shows the time of symptom onset. The horizontal dashed line shows the limit of quantification and the y-axis shows limit of detection reported by Wölfel et al. Inset text shows posterior median and 95% credible intervals of the fitted parameters.

**Fig. S11.**
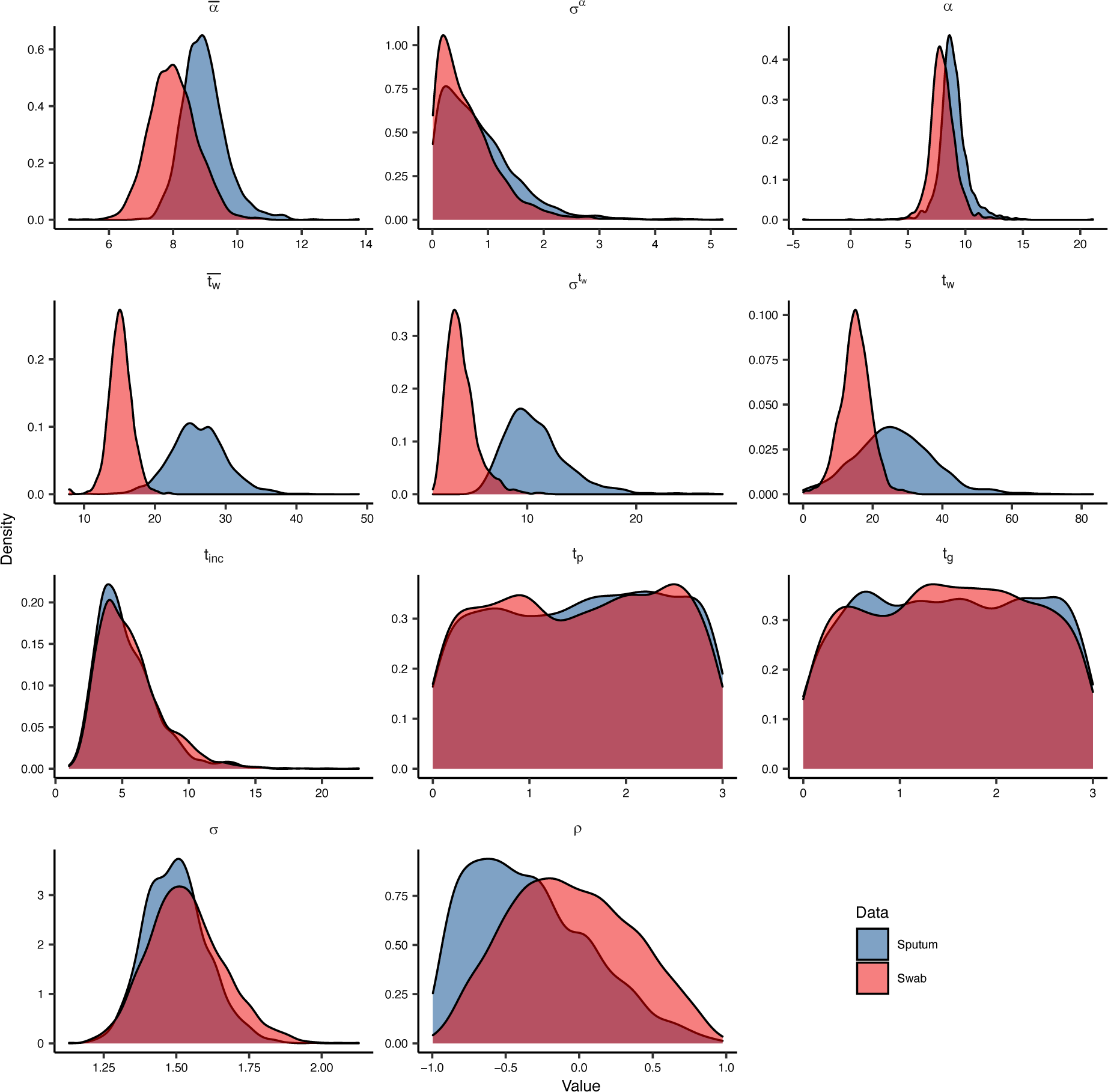
Posterior distributions of estimated parameters fitted to swab and sputum data. Parameters are described in table S5.

**Fig. S12.**
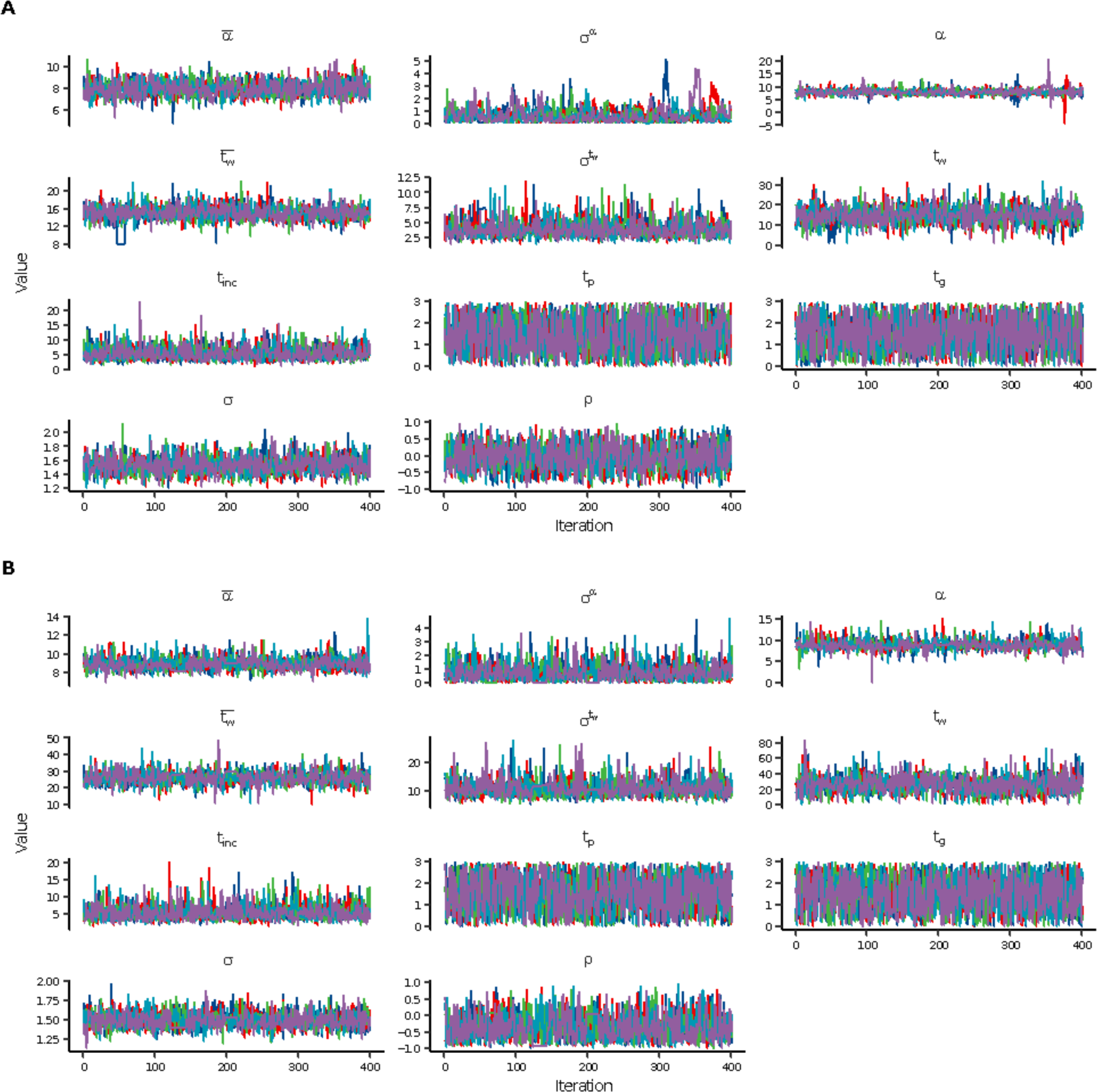
Markov chain Monte Carlo trace plots from fitting to swab and sputum data. Each color shows an independent chain. Trace plots showing the path of the Markov chain Monte Carlo sampler, demonstrating convergence to the same stationary distribution for all chains. Parameters are described in table S5. (A) Swab data. (B) Sputum data.

**Fig. S13.**
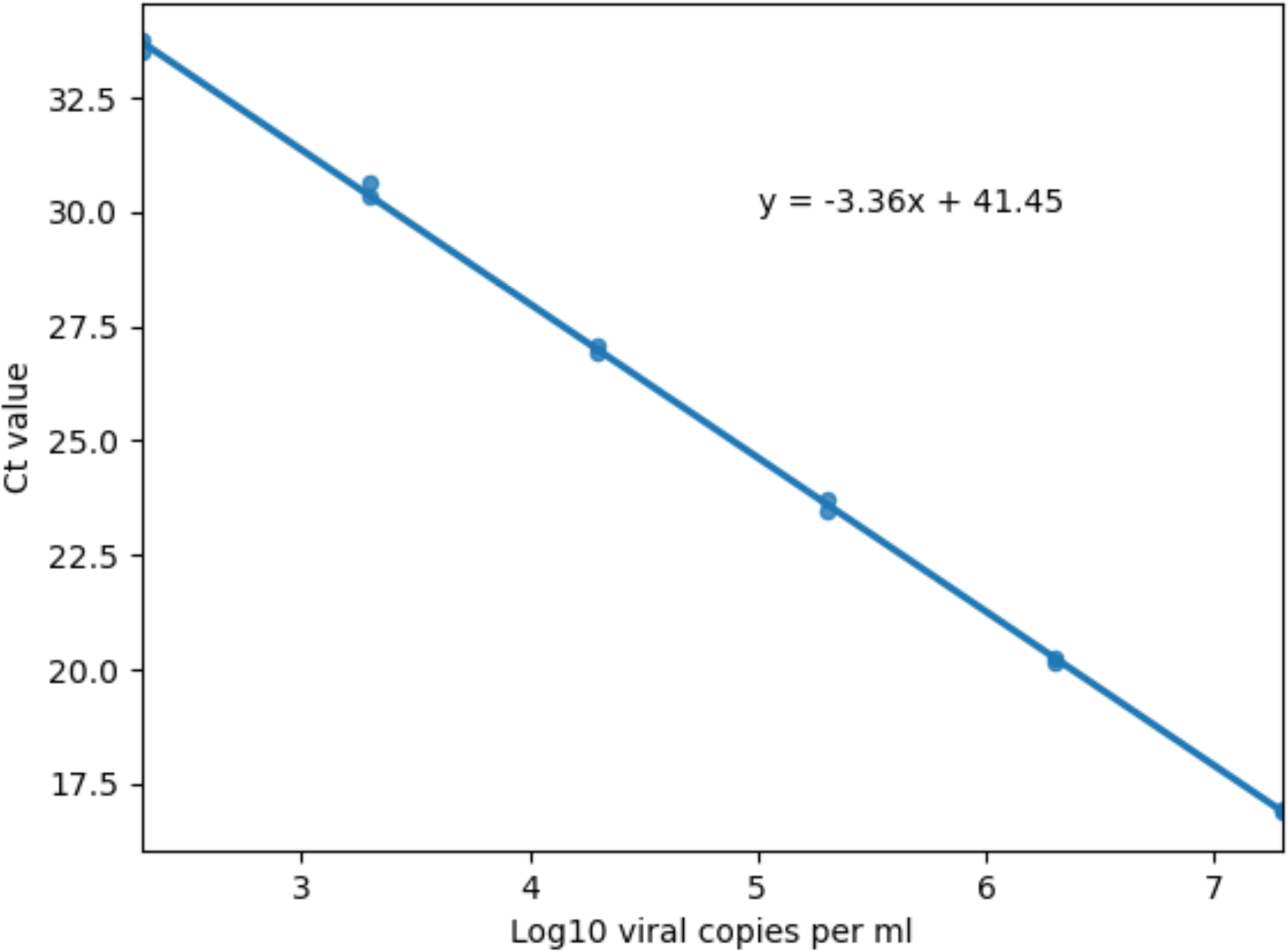
qPCR calibration curve using standard viral RNA copies. qPCR was performed on a dilution series of synthetic Covid RNA with known viral copy numbers (x-axis; log10 copy numbers per ml) to identify Ct values (y-axis) obtained at each copy number. Ct values from replicates at each copy number are indicated by individual points. A linear regression was fit to the data, to establish slope and intercept (equation in panel).

**Table S1.** List of all group test designs for sample identification.

| Number of pools | Number of samples | Number of pools per sample |
| --- | --- | --- |
| 1 | 2 | 1 |
| 1 | 4 | 1 |
| 1 | 8 | 1 |
| 1 | 16 | 1 |
| 1 | 32 | 1 |
| 1 | 64 | 1 |
| 6 | 12 | 2 |
| 6 | 12 | 3 |
| 6 | 24 | 2 |
| 6 | 24 | 3 |
| 6 | 48 | 2 |
| 6 | 48 | 3 |
| 6 | 96 | 2 |
| 6 | 96 | 3 |
| 6 | 192 | 2 |
| 6 | 192 | 3 |
| 6 | 384 | 2 |
| 6 | 384 | 3 |
| 12 | 24 | 2 |
| 12 | 24 | 3 |
| 12 | 24 | 4 |
| 12 | 48 | 2 |
| 12 | 48 | 3 |
| 12 | 48 | 4 |
| 12 | 96 | 2 |
| 12 | 96 | 3 |
| 12 | 96 | 4 |
| 12 | 192 | 2 |
| 12 | 192 | 3 |
| 12 | 192 | 4 |
| 12 | 384 | 2 |
| 12 | 384 | 3 |
| 12 | 384 | 4 |
| 12 | 768 | 2 |
| 12 | 768 | 3 |
| 12 | 768 | 4 |
| 24 | 48 | 2 |
| 24 | 48 | 3 |
| 24 | 48 | 4 |
| 24 | 96 | 2 |
| 24 | 96 | 3 |
| 24 | 96 | 4 |
| 24 | 192 | 2 |
| 24 | 192 | 3 |
| 24 | 192 | 4 |
| 24 | 384 | 2 |
| 24 | 384 | 3 |
| 24 | 384 | 4 |
| 24 | 768 | 2 |
| 24 | 768 | 3 |
| 24 | 768 | 4 |
| 24 | 1536 | 2 |
| 24 | 1536 | 3 |
| 24 | 1536 | 4 |
| 48 | 96 | 2 |
| 48 | 96 | 3 |
| 48 | 96 | 4 |
| 48 | 192 | 2 |
| 48 | 192 | 3 |
| 48 | 192 | 4 |
| 48 | 384 | 2 |
| 48 | 384 | 3 |
| 48 | 384 | 4 |
| 48 | 768 | 2 |
| 48 | 768 | 3 |
| 48 | 768 | 4 |
| 48 | 1536 | 2 |
| 48 | 1536 | 3 |
| 48 | 1536 | 4 |
| 48 | 3072 | 2 |
| **48** | 3072 | 3 |
| **48** | 3072 | 4 |
| **96** | 192 | 2 |
| **96** | 192 | 3 |
| **96** | 192 | 4 |
| **96** | 384 | 2 |
| **96** | 384 | 3 |
| **96** | 384 | 4 |
| **96** | 768 | 2 |
| **96** | 768 | 3 |
| **96** | 768 | 4 |
| **96** | 1536 | 2 |
| **96** | 1536 | 3 |
| **96** | 1536 | 4 |
| **96** | 3072 | 2 |
| **96** | 3072 | 3 |
| **96** | 3072 | 4 |
| **96** | 6144 | 2 |
| **96** | 6144 | 3 |
| **96** | 6144 | 4 |

**Table S2.** Cycle threshold values from qPCR on pooled samples with variable viral load. Five positive samples with variable viral loads (89,000, 12,300, 1,280, 140, and 11, for samples 1-5, respectively) were each pooled with 23 negative samples (“Dil_1” to “Dil_5”). Values in bottom table show the Ct values for each of five pools for three primer pairs (N1, N2, and RP), along with a final call (positive, negative, or inconclusive).

| Sample Name | N1 CT | N2 CT | RP CT | Call |
| --- | --- | --- | --- | --- |
| 24_Dil_1 | 28.93 | 29.58 | 28.62 | POS |
| 24_Dil_2 | 31.07 | 31.73 | 28.71 | POS |
| 24_Dil_3 | 33.29 | 33.91 | 28.37 | POS |
| 24_Dil_4 | Undetermined | 35.9 | 29.17 | Inconclusive |
| 24_Dil_5 | Undetermined | Undetermined | 28.62 | Negative |
| Pool_1 | 30.49 | 31.18 | 28.3 | POS |
| Pool_2 | 31.49 | 31.93 | 28.75 | POS |
| Pool_3 | Undetermined | Undetermined | 29.62 | Negative |
| Pool_4 | 31.71 | 31.66 | 29.34 | POS |
| Pool_5 | Undetermined | Undetermined | 30.01 | Negative |
| Pool_6 | Undetermined | Undetermined | 29.99 | Negative |

**Table S3.**
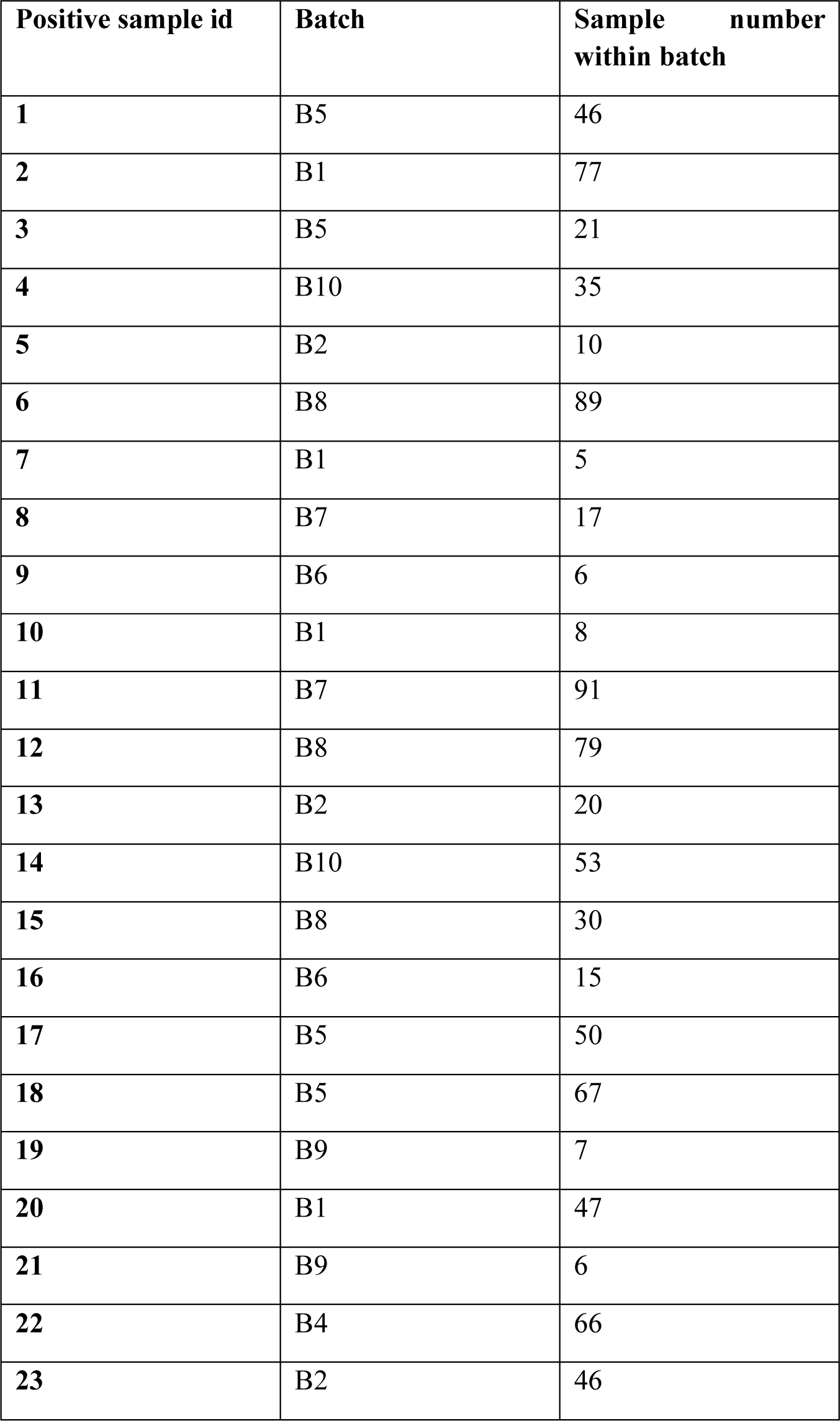
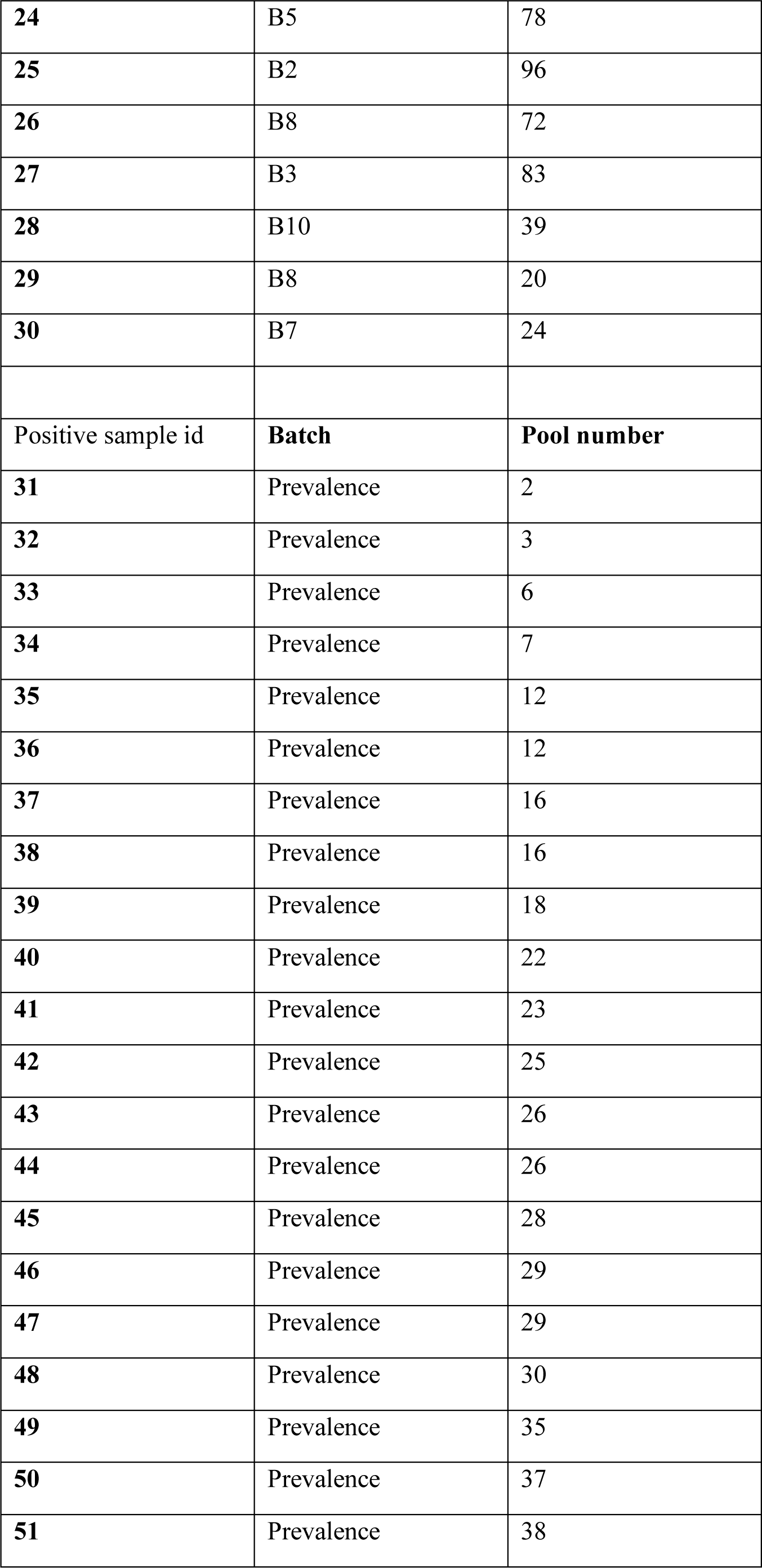
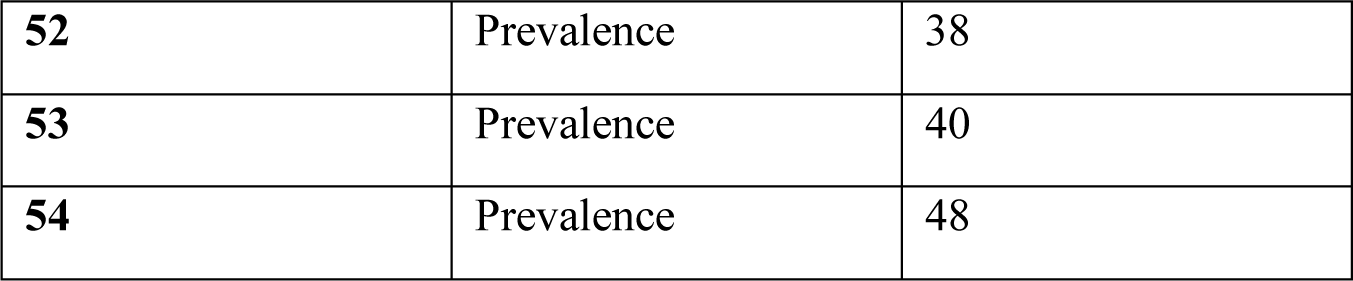
Positive sample distribution within validation pools. 30 positive samples were randomly distributed across batches of combinatorial pooling (B1-B10) and randomly assigned sample numbers within batches. 24 separate positive samples were randomly assigned to 1 of 48 simple pools (“Prevalence”, pool number 2…48).

**Table S4.** Pool design for combinatorial test with 96 samples. Each of 96 samples was split in equal volume into 2 out of 6 pools.

| Sample | Pools |  |
| --- | --- | --- |
| 1 | A | B |
| 2 | A | B |
| 3 | A | B |
| 4 | A | B |
| 5 | A | B |
| 6 | A | B |
| 7 | A | B |
| 8 | C | D |
| 9 | C | D |
| 10 | C | D |
| 11 | C | D |
| 12 | C | D |
| 13 | C | D |
| 14 | C | D |
| 15 | E | F |
| 16 | E | F |
| 17 | E | F |
| 18 | E | F |
| 19 | E | F |
| 20 | E | F |
| 21 | E | F |
| 22 | B | C |
| 23 | B | C |
| 24 | B | C |
| 25 | B | C |
| 26 | B | C |
| 27 | B | C |
| 28 | B | C |
| 29 | D | F |
| 30 | D | F |
| 31 | D | F |
| 32 | D | F |
| 33 | D | F |
| 34 | D | F |
| 35 | D | F |
| 36 | A | E |
| 37 | A | E |
| 38 | A | E |
| 39 | A | E |
| 40 | A | E |
| 41 | A | E |
| 42 | A | E |
| 43 | B | D |
| 44 | B | D |
| 45 | B | D |
| 46 | B | D |
| 47 | B | D |
| 48 | B | D |
| 49 | A | F |
| 50 | A | F |
| 51 | A | F |
| 52 | A | F |
| 53 | A | F |
| 54 | A | F |
| 55 | C | E |
| 56 | C | E |
| 57 | C | E |
| 58 | C | E |
| 59 | C | E |
| 60 | C | E |
| 61 | B | E |
| 62 | B | E |
| 63 | B | E |
| 64 | B | E |
| 65 | B | E |
| 66 | B | E |
| 67 | C | F |
| 68 | C | F |
| 69 | C | F |
| 70 | C | F |
| 71 | C | F |
| 72 | C | F |
| 73 | A | D |
| 74 | A | D |
| 75 | A | D |
| 76 | A | D |
| 77 | A | D |
| 78 | A | D |
| 79 | B | F |
| 80 | B | F |
| 81 | B | F |
| 82 | B | F |
| 83 | B | F |
| 84 | B | F |
| 85 | D | E |
| 86 | D | E |
| 87 | D | E |
| 88 | D | E |
| 89 | D | E |
| 90 | D | E |
| 91 | A | C |
| 92 | A | C |
| 93 | A | C |
| 94 | A | C |
| 95 | A | C |
| 96 | A | C |

**Table S5.** Description of all parameters used in the viral kinetics and transmission models. Values shown are posterior median estimates and 95% credible intervals or fixed value.

| Parameter | Description | Posterior mean (95% CI) | Prior (parameters) |
| --- | --- | --- | --- |
| **Viral kinetics model (swab)** |  |  |  |
| $\bar{\alpha}$ | Mean of peak viral load parameters | 7.98 (6.66-9.47) | Uniform(-100,100) |
| $\sigma_{\alpha}$ | Standard deviation of peak viral load parameters | 0.498 (0.0254-2.13) | Half-Cauchy (1) |
| $\alpha^i$ | Individual peak viral load parameters | Varied | Multivariate normal |
| $\bar{t}_w$ | Mean time from peak viral load to undetectable | 15.1 (12-18.4) | Uniform(-100,100) |
| $\sigma_{t_w}$ | Standard deviation of viral waning parameters | 3.72 (2.02-7.55) | Half-Cauchy (1) |
| $t_w$ | Individual time from peak viral load to undetectable | Varied | Multivariate normal |
| $t_{inc}$ | Time from infection to symptom onset | 5.52 (2.23-11.5) | Log normal (1.621, 0.418) |
| $t_g$ | Duration of pre-viral growth latent period | Varied | Uniform(0,3) |
| $t_p$ | Time from growth initiation to peak viral load | Varied | Uniform(0,3) |
| $\sigma_{obs}$ | Standard deviation of observation process | 1.52 (1.31-1.82) | Uniform(0,10) |
| $\rho_{\alpha, t_w}$ | Correlation between $\bar{\alpha}$ and $\bar{t}_w$ | -0.0808 (-0.79-0.765) | LKJ correlation matrix ($\eta=2$) |
| **Viral kinetics model (sputum)** |  |  |  |
| $\bar{\alpha}$ | Mean of peak viral load parameters | 8.91 (7.8-10.5) | Uniform(-100,100) |
| $\sigma_{\alpha}$ | Standard deviation of peak viral load parameters | 0.674 (0.0417-2.32) | Half-Cauchy (1) |
| $\alpha^i$ | Individual peak viral load parameters | Varied | Multivariate normal |
| $\bar{t}_w$ | Mean time from peak viral load to undetectable | 26.0 (18.2-34.4) | Uniform(-100,100) |
| $\sigma_{t_w}$ | Standard deviation of viral waning parameters | 10.4 (6.51-18.2) | Half-Cauchy (1) |
| $t_w$ | Individual time from peak viral load to undetectable | Varied | Multivariate normal |
| $t_{inc}$ | Time from infection to symptom onset | 5.52 (2.23-11.5) | Log normal (1.621, 0.418) |
| $t_g$ | Duration of pre-viral growth latent period | Varied | Uniform(0,3) |
| $t_p$ | Time from growth initiation to peak viral load | Varied | Uniform(0,3) |
| $\sigma_{obs}$ | Standard deviation of observation process | 1.49 (1.31-1.73) | Uniform(0,10) |
| $\rho_{\alpha, t_w}$ | Correlation between $\bar{\alpha}$ and $\bar{t}_w$ | -0.398 (-0.925-0.579) | LKJ correlation matrix ($\eta=2$) |
| **SEIR Model** |  |  |  |
| $R_0$ | Basic reproductive number (main analyses) | 2.5 (fixed) | NA |
| $R_0^2$ | Basic reproductive number after day 80 (multiple wave epidemic analyses) | 0.8 (fixed) | NA |
| $R_0^3$ | Basic reproductive number after day 150 (multiple wave epidemic analyses) | 1.5 (fixed) | NA |
| $1/\gamma$ | Infectious period | 7 days (fixed) | NA |
| $1/\sigma$ | Incubation period | 6.4 days (fixed) | NA |
| $I_0$ | Initial number of infected individuals | 100 (fixed) | NA |
| $N$ | Total population size | 12500000 (fixed) | NA |
| $R_0$ | Basic reproductive number (main analyses) | 2.5 (fixed) | NA |

**Table S6:**
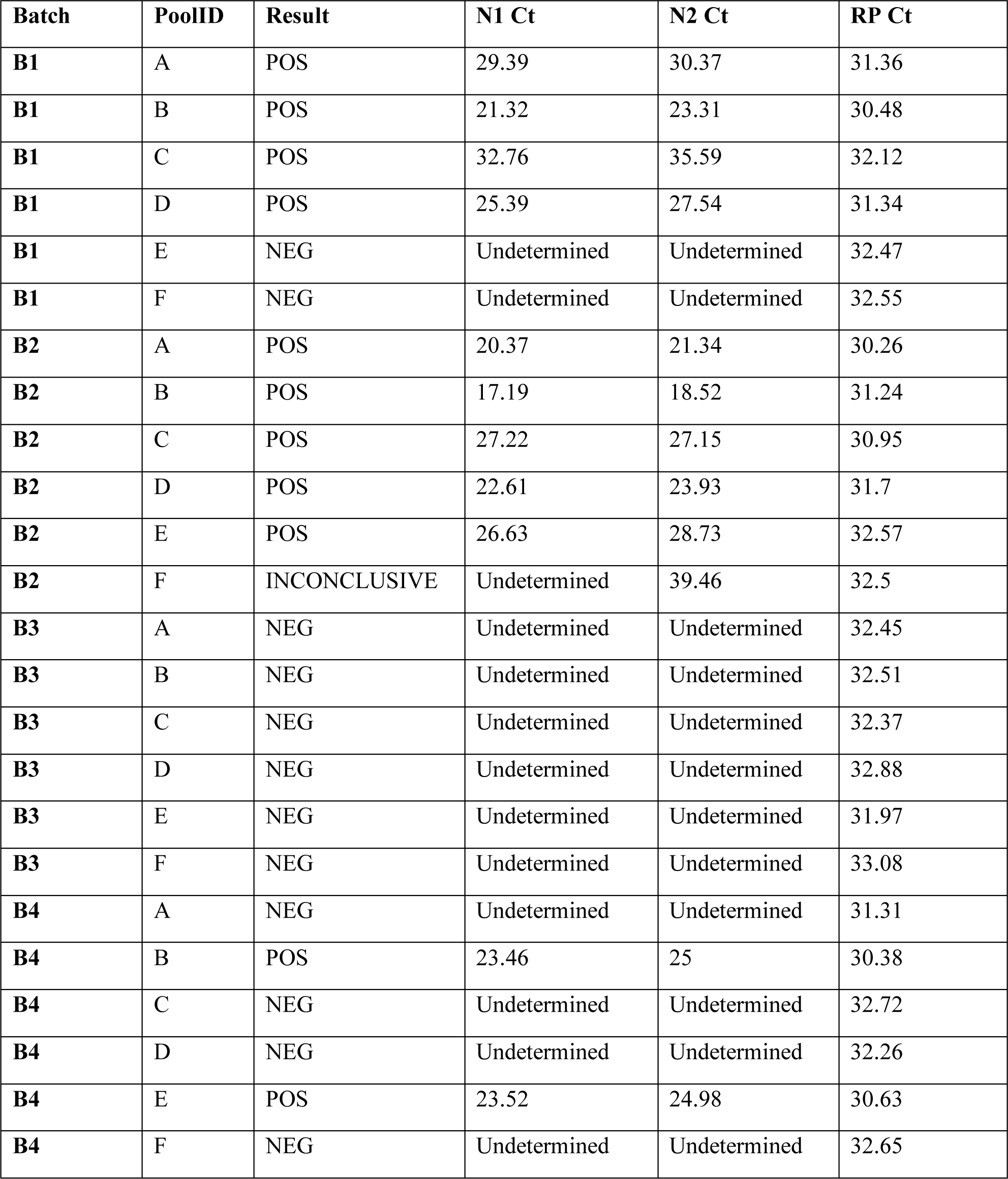
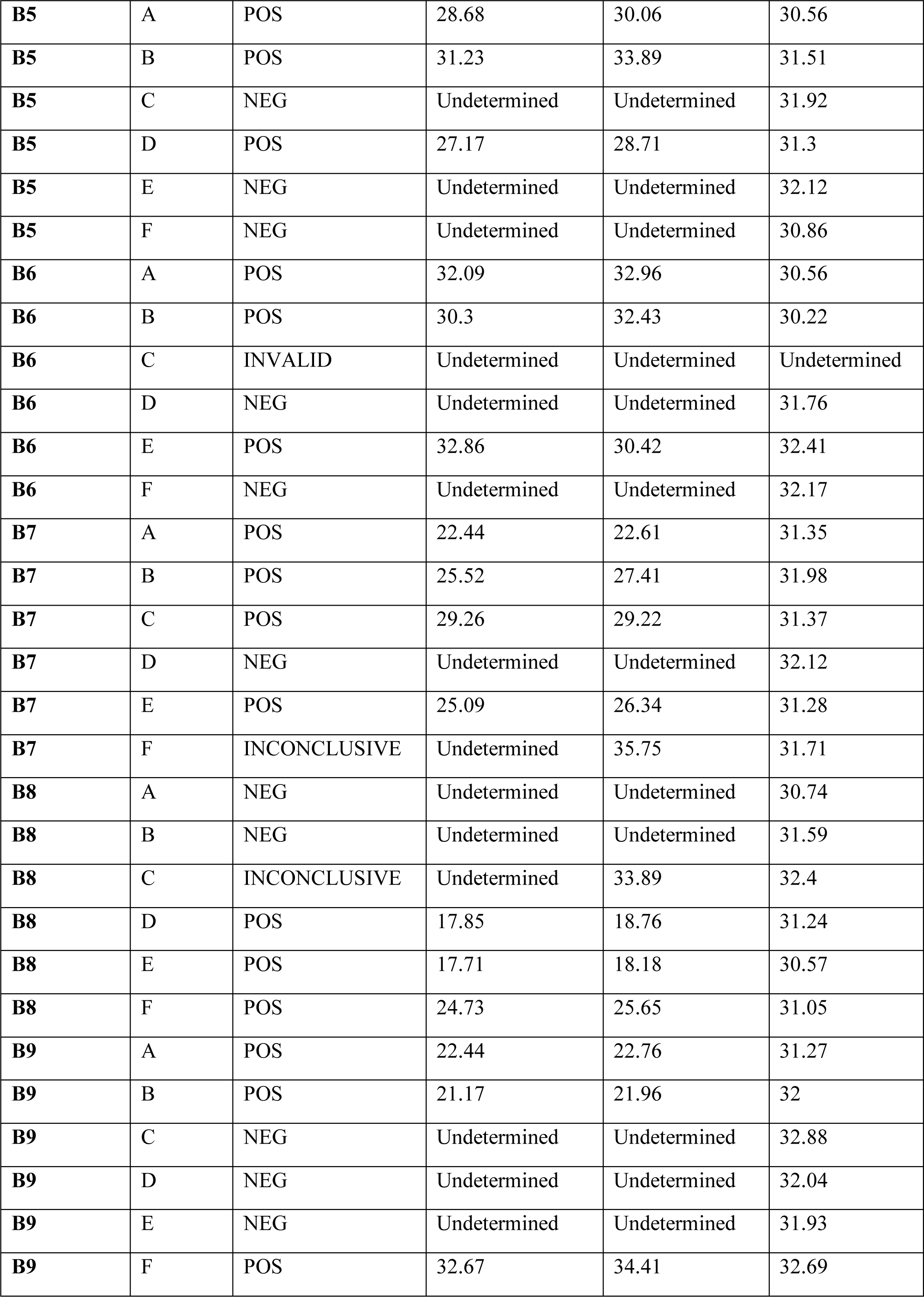
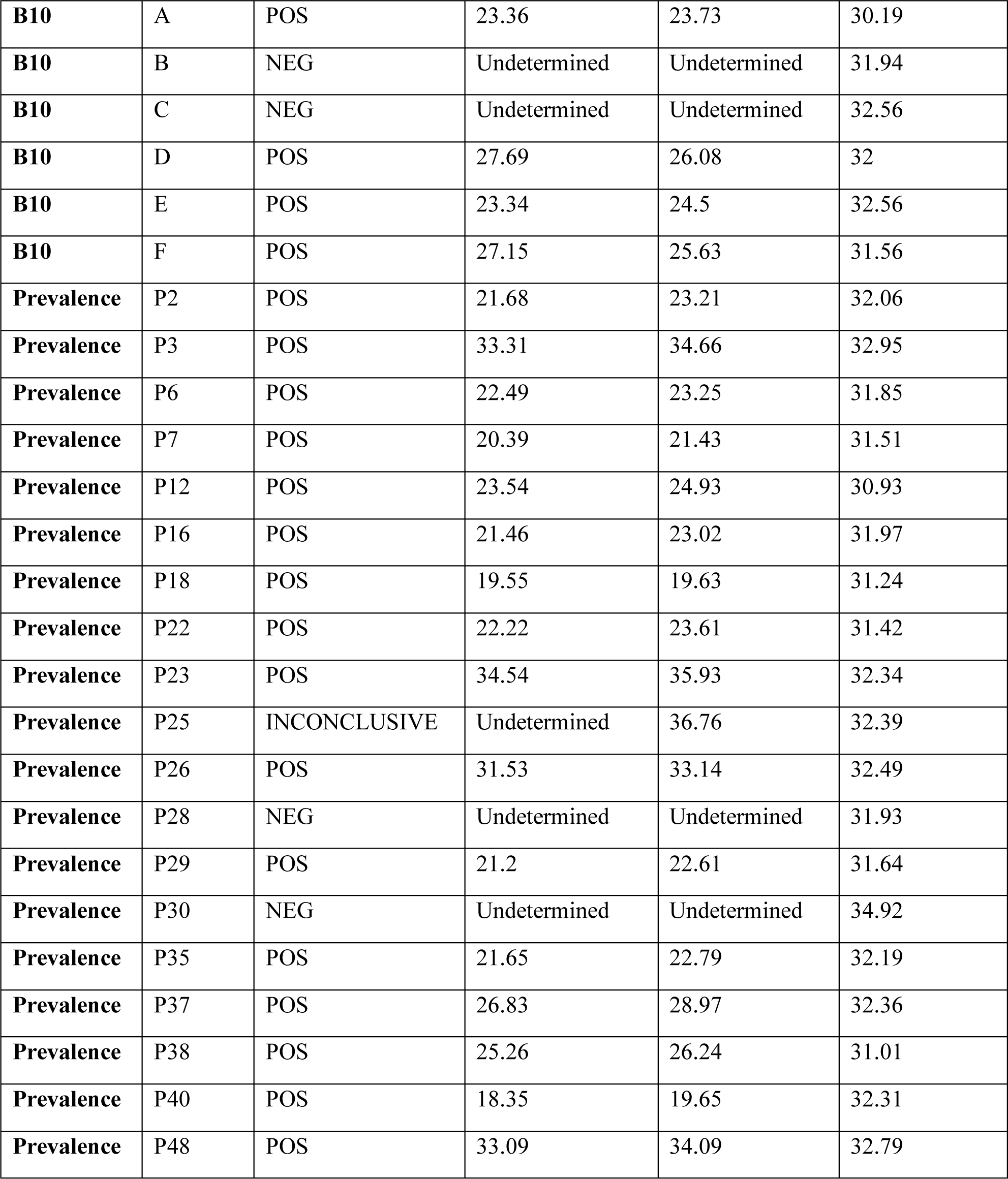
RT-qPCR results for pooling validations. 10 batches of combinatorial pools (B1-B10, pools A-F in each batch) and 19 simple pools (“Prevalence”, pools P2…P48) were tested by RT-qPCR. Test results (POS, NEG, INCONCLUSIVE, or INVALID) and Ct values for N1, N2, and RP primers are shown for each pool.

| **Batch** | **PoolID** | **Result** | **N1 Ct** | **N2 Ct** | **RP Ct** |
| --- | --- | --- | --- | --- | --- |
| **B1** | A | POS | 29.39 | 30.37 | 31.36 |
| **B1** | B | POS | 21.32 | 23.31 | 30.48 |
| **B1** | C | POS | 32.76 | 35.59 | 32.12 |
| **B1** | D | POS | 25.39 | 27.54 | 31.34 |
| **B1** | E | NEG | Undetermined | Undetermined | 32.47 |
| **B1** | F | NEG | Undetermined | Undetermined | 32.55 |
| **B2** | A | POS | 20.37 | 21.34 | 30.26 |
| **B2** | B | POS | 17.19 | 18.52 | 31.24 |
| **B2** | C | POS | 27.22 | 27.15 | 30.95 |
| **B2** | D | POS | 22.61 | 23.93 | 31.7 |
| **B2** | E | POS | 26.63 | 28.73 | 32.57 |
| **B2** | F | INCONCLUSIVE | Undetermined | 39.46 | 32.5 |
| **B3** | A | NEG | Undetermined | Undetermined | 32.45 |
| **B3** | B | NEG | Undetermined | Undetermined | 32.51 |
| **B3** | C | NEG | Undetermined | Undetermined | 32.37 |
| **B3** | D | NEG | Undetermined | Undetermined | 32.88 |
| **B3** | E | NEG | Undetermined | Undetermined | 31.97 |
| **B3** | F | NEG | Undetermined | Undetermined | 33.08 |
| **B4** | A | NEG | Undetermined | Undetermined | 31.31 |
| **B4** | B | POS | 23.46 | 25 | 30.38 |
| **B4** | C | NEG | Undetermined | Undetermined | 32.72 |
| **B4** | D | NEG | Undetermined | Undetermined | 32.26 |
| **B4** | E | POS | 23.52 | 24.98 | 30.63 |
| **B4** | F | NEG | Undetermined | Undetermined | 32.65 |
| **B5** | A | POS | 28.68 | 30.06 | 30.56 |
| **B5** | B | POS | 31.23 | 33.89 | 31.51 |
| **B5** | C | NEG | Undetermined | Undetermined | 31.92 |
| **B5** | D | POS | 27.17 | 28.71 | 31.3 |
| **B5** | E | NEG | Undetermined | Undetermined | 32.12 |
| **B5** | F | NEG | Undetermined | Undetermined | 30.86 |
| **B6** | A | POS | 32.09 | 32.96 | 30.56 |
| **B6** | B | POS | 30.3 | 32.43 | 30.22 |
| **B6** | C | INVALID | Undetermined | Undetermined | Undetermined |
| **B6** | D | NEG | Undetermined | Undetermined | 31.76 |
| **B6** | E | POS | 32.86 | 30.42 | 32.41 |
| **B6** | F | NEG | Undetermined | Undetermined | 32.17 |
| **B7** | A | POS | 22.44 | 22.61 | 31.35 |
| **B7** | B | POS | 25.52 | 27.41 | 31.98 |
| **B7** | C | POS | 29.26 | 29.22 | 31.37 |
| **B7** | D | NEG | Undetermined | Undetermined | 32.12 |
| **B7** | E | POS | 25.09 | 26.34 | 31.28 |
| **B7** | F | INCONCLUSIVE | Undetermined | 35.75 | 31.71 |
| **B8** | A | NEG | Undetermined | Undetermined | 30.74 |
| **B8** | B | NEG | Undetermined | Undetermined | 31.59 |
| **B8** | C | INCONCLUSIVE | Undetermined | 33.89 | 32.4 |
| **B8** | D | POS | 17.85 | 18.76 | 31.24 |
| **B8** | E | POS | 17.71 | 18.18 | 30.57 |
| **B8** | F | POS | 24.73 | 25.65 | 31.05 |
| **B9** | A | POS | 22.44 | 22.76 | 31.27 |
| **B9** | B | POS | 21.17 | 21.96 | 32 |
| **B9** | C | NEG | Undetermined | Undetermined | 32.88 |
| **B9** | D | NEG | Undetermined | Undetermined | 32.04 |
| **B9** | E | NEG | Undetermined | Undetermined | 31.93 |
| **B9** | F | POS | 32.67 | 34.41 | 32.69 |
| **B10** | A | POS | 23.36 | 23.73 | 30.19 |
| **B10** | B | NEG | Undetermined | Undetermined | 31.94 |
| **B10** | C | NEG | Undetermined | Undetermined | 32.56 |
| **B10** | D | POS | 27.69 | 26.08 | 32 |
| **B10** | E | POS | 23.34 | 24.5 | 32.56 |
| **B10** | F | POS | 27.15 | 25.63 | 31.56 |
| **Prevalence** | P2 | POS | 21.68 | 23.21 | 32.06 |
| **Prevalence** | P3 | POS | 33.31 | 34.66 | 32.95 |
| **Prevalence** | P6 | POS | 22.49 | 23.25 | 31.85 |
| **Prevalence** | P7 | POS | 20.39 | 21.43 | 31.51 |
| **Prevalence** | P12 | POS | 23.54 | 24.93 | 30.93 |
| **Prevalence** | P16 | POS | 21.46 | 23.02 | 31.97 |
| **Prevalence** | P18 | POS | 19.55 | 19.63 | 31.24 |
| **Prevalence** | P22 | POS | 22.22 | 23.61 | 31.42 |
| **Prevalence** | P23 | POS | 34.54 | 35.93 | 32.34 |
| **Prevalence** | P25 | INCONCLUSIVE | Undetermined | 36.76 | 32.39 |
| **Prevalence** | P26 | POS | 31.53 | 33.14 | 32.49 |
| **Prevalence** | P28 | NEG | Undetermined | Undetermined | 31.93 |
| **Prevalence** | P29 | POS | 21.2 | 22.61 | 31.64 |
| **Prevalence** | P30 | NEG | Undetermined | Undetermined | 34.92 |
| **Prevalence** | P35 | POS | 21.65 | 22.79 | 32.19 |
| **Prevalence** | P37 | POS | 26.83 | 28.97 | 32.36 |
| **Prevalence** | P38 | POS | 25.26 | 26.24 | 31.01 |
| **Prevalence** | P40 | POS | 18.35 | 19.65 | 32.31 |
| **Prevalence** | P48 | POS | 33.09 | 34.09 | 32.79 |

